# Age and CMV are not associated with clinical or immunological response to immune checkpoint blockade

**DOI:** 10.64898/2026.08.04.26359748

**Authors:** Jack M Edwards, Sashendra Senthi, Robin Smith, Hayley Burridge, Carole Owens, Mark Shackleton, Miles C Andrews, Menno C van Zelm

**Affiliations:** Allergy and Clinical Immunology Laboratory, Department of Immunology, School of Translational Medicine, Monash University and Alfred Hospital, Melbourne, VIC, Australia; Cancer and Host Dynamics Group, Department of Cancer Medicine, School of Translational Medicine, Monash University, Melbourne, VIC, Australia; Alfred Health Radiation Oncology, The Alfred Hospital, Melbourne, VIC, Australia; Department of Medical Oncology, The Alfred Hospital, Melbourne, VIC, Australia; Department of Immunology, Erasmus MC, University Medical Center, Rotterdam, The Netherlands

## Abstract

Ageing and cytomegalovirus (CMV) infection drive major alterations to T-cell immunity. Age is also associated with an increased risk of cancers including melanoma, which is treated with T-cell modifying immune checkpoint blockade (ICB). However, the extent to which age, CMV, and treatment-induced immune changes interact to shape clinical outcomes remains poorly understood. We investigated this through flow cytometric evaluation of pre- and early on-treatment blood samples of 79 advanced melanoma patients. Age and CMV infection were associated with significant and largely distinct changes to T cell phenotype pre-treatment but had no impact on clinical outcome. Older patients (≥65 years) had fewer CD8^+^ Tnaive, CD4^+^ Tcm, TFH, and B cells, and increased CD8^+^ TemRA, but similar cytokine and inhibitory marker expression. Conversely, CMV drove expansion of CD8^+^ and CD4^+^ TemRA cells with enhanced effector function without reducing naive populations. One cycle of PD-1 and CTLA-4 ICB induced immune cell expansion and phenotype changes of greater magnitude and partially distinct from those seen during PD-1 with or without LAG-3 ICB, but these effects were largely independent of age or CMV serostatus. Hence, neither ageing nor CMV were associated with clinical outcome or immunological response to ICB in advanced melanoma patients.

## Introduction

The incidence of cancer increases with age, with over half of all cancer diagnoses occurring over the age of 65 (1–3). Concordantly, ageing is associated with significant molecular changes to the body, with major alterations occurring around 45 years and 60-65 years of age (4). These changes extend to the innate and adaptive immune responses, which change dynamically over the course of life in a process termed immune ageing (3,4). Immune ageing is associated with thymic involution and immune senescence, processes which lead to decreases in circulating naive T cells, particularly CD8^+^ cells, and a subsequent loss of T-cell receptor (TCR) diversity and proportional enrichment in terminally differentiated subsets (4,5). Depletion of naive T cells and TCR diversity may reduce the capacity of the aged immune system to recognize and mount responses against tumor neoantigens, while the accumulation of senescent cells leads to a ‘senescence-associated secretory phenotype’ (SASP) whereby cells release pro-inflammatory cytokines, including IL-1, IL-6, IL-8, and TNFα, that contribute to a constant state of low-grade inflammation known as inflammaging (6,7). Higher levels of IL-6, IL-8, and expansion of a senescent CD57^+^ KLRG1^+^ CD28^-^ CD8^+^ T cell subset have been linked with poorer response rates, shorter progression-free survival (PFS), and reduced overall survival (OS) following immune checkpoint blockade (ICB) treatment of solid cancers (8–12). Similarly, naive and memory B-cell abundance declines with age (4), and older patients have impaired antibody responses (13), although these vary greatly between individuals (3). Thus, immune ageing results in decreased immune fitness, potentially impacting immune surveillance and limiting the efficacy of ICB.

Alongside ageing, chronic infection with cytomegalovirus (CMV) has a significant impact on T-cell subset composition and their phenotypes (14,15). CMV infection is lifelong, present in approximately 83% of the global population (16), and leads to an expansion of functional non-exhausted CD8^+^ and CD4^+^ T effector memory (TemRA) cells in a process termed ‘memory inflation’ (17). These expanded cell populations express heightened levels of the cytotoxic effector molecules granzyme B, IFNγ, and TNFα, as well as CD57, TBET and EOMES, but decreased PD-1 (18). Thus, ageing and CMV have partially overlapping impacts on T-cell phenotypes, with both leading to proportional enrichment of effector memory subsets over naive cells.

Crucially, T cells are the key effectors of ICB and anti-tumor immunity, and an increased abundance of effector memory T-cell subsets and enrichment of IFNγ signaling have been associated with improved outcomes following ICB (19–23). Thus, both ageing and CMV infection lead to alterations that may affect responses to ICB. While a recent study found that CMV^+^ patients had improved OS following PD-1 blockade and that CMV infection may be protective against the onset of melanoma (24), these associations are not always evident (25), particularly for combined PD-1 and CTLA-4 blockade (24,26). Conversely, and despite preclinical evidence in mouse models suggesting that ICB responsiveness is decreased with age (27–30), retrospective studies in human cohorts have generally shown that age does not diminish objective response rates to ICB or affect OS and PFS (31–40). Indeed, studies within melanoma have shown that ICB in older patients may even outperform treatment in younger patients (32,41), although global immune characteristics may be only one of several factors contributing to this observation.

Given that clinical response is not consistently found to be impacted by age or CMV status, and that on-treatment immune features are more strongly associated with clinical outcomes than pre-treatment features (23,42,43), we hypothesized that both clinical outcome and dynamic immunological responses to ICB would be maintained in patients stratified by age or CMV status. To address this question, we collected peripheral blood samples from patients undergoing treatment of advanced melanoma with PD-1 ± CTLA-4 or LAG-3 blockade and used high-parameter spectral flow cytometry to robustly interrogate innate, B, and T cell immunity prior to commencing and early during treatment. We found that clinical response was maintained regardless of age and CMV serostatus, which each drove distinct changes in baseline immunity but did not affect immunological response to treatment. We further compared the immunological effects of single agent PD-1, PD-1 + CTLA-4, and PD-1 + LAG-3 blockade, and found that PD-1 + CTLA-4 blockade induced significantly larger immune expansion and transformation than PD-1 with or without LAG-3 blockade.

## Materials and methods

### Patients and healthy volunteers

79 patients with unresectable stage III or IV melanoma receiving standard of care ICB at the Alfred Hospital, Melbourne, Australia, were prospectively recruited to the Alfred Cancer Biobank (Alfred Ethics Committee 535/19) for an observational registry-based study. Written informed consent was obtained from all participants prior to enrolment. Cohort sizes were dictated by knowledge of effect sizes from previous work (43) and sample availability. Patients received either PD-1 blockade monotherapy with nivolumab (240mg intravenously 2-weekly or 480mg intravenously 4-weekly) or pembrolizumab (200mg intravenously 3-weekly or 400mg intravenously 6-weekly), combined ipilimumab 3mg/kg + nivolumab 1mg/kg intravenously 3-weekly for up to four cycles, followed by maintenance nivolumab 3mg/kg or 480mg (flat-dosing) 4-weekly (ipi+nivo), or combined relatlimab + nivolumab fixed-dose combination (480mg nivolumab + 160mg relatlimab-rmbw, intravenously 4-weekly). Blood samples were obtained pre- and early on-treatment (immediately prior to cycle 2), and stored PBMC were retrieved from the biobank under the Translational Research in Immunotherapy Patients study (TRIP; AEC 507/20). Best overall response (BOR) was assessed by treating clinicians using RECIST v1.1 guidelines (44). Immune-related adverse events were based on documented events graded according to the Common Terminology Criteria for Adverse Events (CTCAE) version 5.0 (45) by the treating clinician(s) and/or clinical staff of the Alfred Cancer Biobank. 21 healthy controls were recruited under an approved protocol from Monash University (26385) (43). All studies were conducted in accordance with the ethical principles of the Declaration of Helsinki and with adherence to the Good Clinical Practice guidelines as defined by the International Conference on Harmonization.

### Sample processing

Blood samples were collected in Vacutainer K2EDTA blood tubes (BD Biosciences, San Jose, CA). Total leukocyte counts were determined with the Cell Dyn analyzer (Abbott Core Laboratory, Abbott Park, IL, USA), and flow cytometric TruCount analysis (see below). Blood samples were centrifuged at 1,320 g for 10 min, plasma was removed and centrifuged again at 12,000 g for 10 min before storage at −80°C. The remaining blood was diluted 1:1 with phosphate-buffered saline (PBS) and loaded onto Ficoll-Hypaque Plus (Cytiva, Marlborough, MA, USA) for isolation of peripheral blood mononuclear cells (PBMCs) by density gradient centrifugation at 650 g for 25 min with no brake. After washing twice with PBS, PBMCs were cryopreserved in liquid nitrogen at a density of 10 million viable cells/mL in 50% FCS (Bovogen, Melbourne, VIC, Australia), 40% RPMI-1640 (Sigma-Aldrich, St. Louis, MO, USA), and 10% DMSO (Sigma-Aldrich).

### TruCount analysis

Absolute numbers of leukocytes, granulocytes, monocytes, lymphocytes, and subsets therein were determined using a lyse-no-wash T, B and NK TruCount assay, as previously described (46). Briefly, within 6 h of sample collection, 50 μL of whole blood was added to a TruCount tube (BD Biosciences) together with an antibody cocktail of 20 μL containing conjugated primary antibodies against human CD3, CD4, CD8, CD14, CD16, CD19, CD45, CD56 and HLA-DR (**Supplementary Table 1**). Following incubation for 15 min at room temperature, samples were lysed with FACS Lysing Solution (BD Biosciences) in a total volume of 500 μL for 15 min and stored at 4°C in the dark for up to 1 hour before acquisition on a Fortessa LSRII or FACSLyric flow cytometer (BD Biosciences).

### T cell flow cytometry

Cryopreserved PBMCs were batch-thawed and stained using two previously-described spectral flow cytometry panels for in-depth phenotyping of T cells (46). One tube was designed to interrogate surface and intracellular marker expression by resting *ex vivo* T cells, and a second to evaluate the production of effector molecules after overnight stimulation with CD3 and CD28 in the presence of Brefeldin A and Monensin. Panel composition and antibody details are listed in **Supplementary Table 2**. Samples were acquired on a 5-laser Cytek Aurora with standard Cytek Assay Settings adjusted daily using SpectroFlo Quality Control (QC) beads (Cytek Biosciences, Fremont, CA, USA) as per the manufacturer’s recommendations. FSC and SSC were adjusted to optimally identify the lymphocyte population, and the FSC area scaling factor was set to 0.95. Samples were acquired using the live unmixing functionality and run at a medium flow rate, averaging 3000–5000 events/s. A median of 632,417 viable T cells were assessed per patient per timepoint across the two spectral flow cytometry panels.

### B cell flow cytometry

Cryopreserved PBMCs were batch-thawed as previously described (46) and stained using a 20-parameter spectral flow cytometry panel for detailed evaluation of the memory B cell compartment (47). In addition to antibodies against surface markers, the staining included fluorescent tetramers of the hemagglutinin (HA) protein from the influenza A/Michigan/45/2015 (H1N1) pdm09-like strain for antigen-specific B cell phenotyping (**Supplementary Table 3**). Recombinant HA was produced, purified, biotinylated, and tetramerized as previously described (48). 12.5 million thawed PBMC were incubated for 20 minutes with Live/Dead Blue Fixable Viability Stain in PBS, washed with FACS buffer (0.1% sodium azide and 0.2% bovine serum albumin in PBS), and then stained for 5 minutes with 5 µg/mL of each fluorescent tetramer before addition of remaining antibodies for 15 minutes in a total staining volume of 250 µL. Brilliant stain buffer (BD Biosciences) was included with both tetramers and antibodies. In a separate tube, 2 million PBMC were stained with Live/Dead Blue Fixable Viability stain, fluorescent streptavidin controls without Flu protein, and antibodies against CD3, CD19, CD27, CD38 and IgD as for the other tube in a total volume of 100 µL. Cells were then washed with FACS buffer, fixed with 2% paraformaldehyde (PFA) for 20 minutes, washed twice more, and acquired on a 5-laser Cytek Aurora as above. All incubations were performed at room temperature in the dark. A median of 298,227 viable B cells were assessed per patient, per timepoint.

### Assessment of CMV serostatus

Cytomegalovirus (CMV) seropositivity was assessed by measurement of capsid-specific IgG in patient plasma by ELISA as per the manufacturer’s instructions (Euroimmun AG, Germany). The concentration of CMV-specific IgG was interpolated from a standard curve generated using the provided calibration controls and seropositivity defined as ≥22 RU/mL IgG.

### Data analysis

All samples were analyzed using FlowJo v10.10 (BD Biosciences), with minor compensation adjustments performed as required after fluorochrome-specific biexponential scaling. Only samples with high cell viability (>80%) and stability (assessed by tracking CD3 BUV805 or CD19 BUV563 intensity versus time for T and B cell panels, respectively) were included in this study. Manual identification of immune cell populations was performed as previously described for each of the TruCount and T cell panels (**Supplementary Tables 4-6, and Supplementary Figs. 1-3**) (43). B cells were gated as shown in **Supplementary Fig 4** and as per **Supplementary Table 7**. A minimum of 30 tetramer positive events were assessed per sample; samples with fewer events were removed from subsequent phenotype analysis. Using the absolute cell numbers obtained from the TruCount panel, T- and B-cell subset frequencies were converted into absolute counts per µL of blood; to avoid interdependence of subset proportions, we typically chose to report the absolute count of subsets and frequencies of marker expression therein. Data were analyzed and plotted using GraphPad Prism v10.4.1, and the following packages in the R v4.5.1 programming environment: tidyverse v2.0.0, ggpubr v0.6.1, ggplot2 v3.5.2, survival v3.8-3, and vegan v2.7-2 (49–53).

### Statistical analysis

Non-parametric statistical analyses were performed using the Mann-Whitney test, Kruskal-Wallis test, or Wilcoxon signed-rank test as appropriate. Considering the hierarchical, dependent nature of the data, multiple comparisons adjustment was performed using the Bonferroni method with respect to the three major lineages analyzed: CD8^+^ T cells, CD4^+^ T cells, and B cells, leaving an adjusted significance threshold (alpha) for all tests of 0.017. Differences observed at the lineage level were then explored in the dependent subsets within. Graphpad Prism and R packages were used to perform the above non-parametric statistical tests, as well as Log-rank (Mantel-Cox) survival analysis, Spearman correlation, multiple logistic and linear regression, Cox Proportional Hazard’s Regression, and Fisher’s exact tests. Principal component analysis (PCA), Bray-Curtis dissimilarity (9,999 permutations), PERMANOVA, and PERMDISP were performed using the R programming environment. Three samples (one pre-treatment ipi+nivo, one on-treatment ipi+nivo, and one on-treatment nivo/pembro patient) were missing data from the activated T cell panel and were excluded from these analyses. P-values <0.017 were considered significant; p-values greater than this but below 0.05 were reported as trends.

## Results

### Patient cohort and clinical outcomes

79 patients with unresectable stage III or IV melanoma scheduled to be treated with immune checkpoint blockade were recruited between November 2020 and May 2025 and paired peripheral blood samples collected prior to and on treatment (**Fig. 1A, Table 1**). 22 patients received PD-1 blockade monotherapy with nivolumab or pembrolizumab (nivo/pembro), 47 received combined CTLA-4 and PD-1 blockade (ipi+nivo), and 10 received combined LAG-3 and PD-1 blockade (rela+nivo). The median age at commencement of treatment was 69 years (range 25-92), with ipi+nivo patients being significantly younger (median 63, range 25-81) than nivo/pembro (76.5, 54-92, *p*=0.0014) and rela+nivo (79, 48-85, *p*=0.0046) patients (**Fig. 1B**). Ipi+nivo patients were also more likely to experience toxicity, have a more advanced stage, elevated lactate dehydrogenase (LDH) levels, and have received previous treatment (**Table 1**). After a median overall follow-up time of 707 days (nivo/pembro 675 days, ipi+nivo 851 days, and rela+nivo 234 days), similar best overall response (BOR) rates were observed between the three cohorts; however, patients receiving ipi+nivo tended to have a longer median overall survival (OS) (median not reached, 33, and 32 months for ipi+nivo, nivo/pembro, and rela+nivo respectively, *p*=0.13). Cox proportional hazards models controlling for age, CMV serostatus, ECOG performance score, and treatment type (reference: nivo/pembro) found ipi+nivo was associated with longer OS (HR 0.28 [0.093-0.84], *p*=0.022) and shorter toxicity-free survival (TFS; HR = 4.19 [1.40-18.07], *p*=0.023), but no change to PFS (**Fig. 1D-E, Supplementary Fig. 5A**). OS was also significantly associated with best overall response (BOR; *p*<0.0001), with any-grade toxicity (*p*=0.0047), and with stage of disease (*p*=0.0004), as expected (**Table 1**).

**Figure 1.**
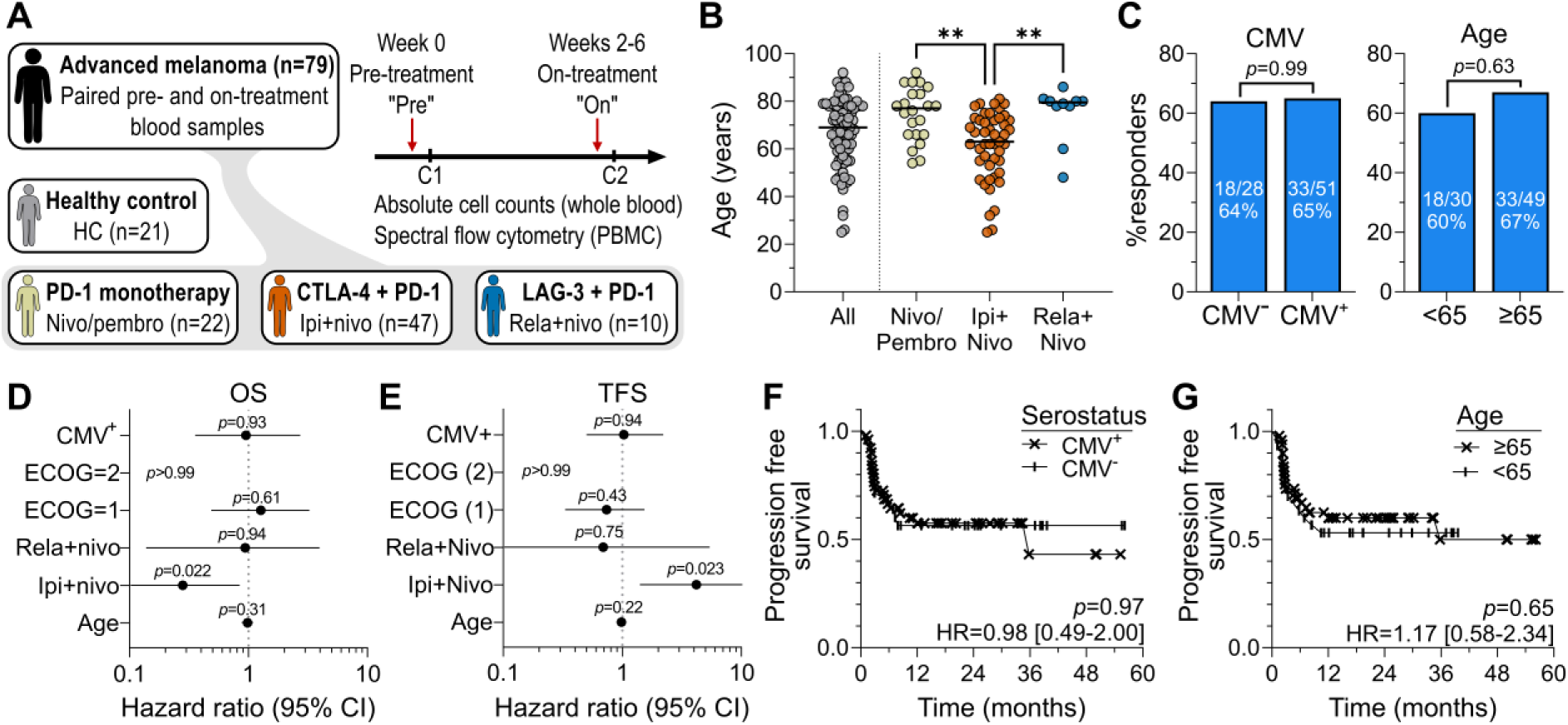
Advanced melanoma patient cohort and clinical outcomes. (**A**) Paired pre- and on-treatment blood samples were collected from advanced melanoma patients treated with single agent PD-1, PD-1 + CTLA-4, or PD-1 + LAG-3, and age-matched healthy controls. (**B**) Age range of all patients and when stratified by treatment; nivo/pembro (n=22), ipi+nivo (n=47), and rela+nivo (n=10). *P* values from Kruskal-Wallis test with Dunn’s Multiple Comparisons test. (**C**) Best overall response rate stratified by CMV serostatus or age. *P* values from Fisher’s exact test. (**D**) Cox proportional hazards model of CMV and age effect on overall survival when controlling for ECOG performance status (reference = 0) and treatment (reference = nivo/pembro). Forest plot shows hazard ratio (HR) and 95% confidence interval (CI). (**E**) Cox proportional hazards as per (D) for toxicity-free survival (TFS). (**F**) Kaplan-Meier progression free survival analysis for CMV status or (**G**) age (±65 years).

**Table 1:** Pre-treatment patient clinical characteristics.

|  | All patients (n=79) | Nivo/Pembro (n=22) | Ipi+Nivo (n=47) | Rela+Nivo (n=10) | p-value<sup>#</sup> (between treatments) | p-value (OS)<sup>§</sup> |
| --- | --- | --- | --- | --- | --- | --- |
| Age (years) |  |  |  |  |  |  |
| Median (range) | 69 (25-92) | 76.5 (54-92) | 63 (25-81) | 79 (48-85) | **0.0001**<sup>^</sup> | 0.53 |
| Sex |  |  |  |  |  |  |
| Male | 57 (72%) | 16 (73%) | 32 (68%) | 9 (90%) | 0.42 | 0.51 |
| Female | 22 (28%) | 6 (27%) | 15 (32%) | 1 (10%) |  |  |
| Best overall response (RECIST v1.1)<sup>‡</sup> |  |  |  |  |  |  |
| Responder (PR, CR) | 51 (65%) | 14 (64%) | 32 (68%) | 5 (50%) | 0.53 | **<0.0001** |
| Non-responder (SD, PD, death) | 28 (35%) | 8 (36%) | 15 (32%) | 5 (50%) |  |  |
| Toxicity (CTCAE v5.0)<sup>‡</sup> |  |  |  |  |  |  |
| Any grade | 65 (82%) | 16 (73%) | 44 (94%) | 5 (50%) | **0.0013** | **0.0047** |
| Grade 3+ | 32 (41%) | 3 (14%) | 28 (60%) | 1 (10%) | **0.0001** | 0.78 |
| Median time (days) to onset of severe toxicity (range) | 47 (14-198) | 50 (42-127) | 47 (14-198) | 18 (NA) | 0.24 | 0.11 |
| Genomics |  |  |  |  |  |  |
| *BRAF* V600 mutant | 19 (24%) | 1 (5%) | 16 (34 %) | 2 (20%) | **0.041** | 0.82 |
| *NRAS* mutant | 29 (38%) | 11 (50%) | 14 (30%) | 4 (40%) |  |  |
| *BRAF/NRAS* wt | 29 (37%) | 8 (36%) | 17 (36%) | 4 (40%) |  |  |
| Not measured | 2 (3%) | 2 (9%) | 0 (0%) | 0 (0%) |  |  |
| Stage of disease (AJCC 8th Ed.)<sup>‡</sup> |  |  |  |  |  |  |
| IIIB | 2 (3%) | 0 (0%) | 0 (0%) | 2 (20%) | **0.0009** | **0.0004** |
| IIIC | 7 (9%) | 3 (14%) | 3 (6%) | 1 (10%) |  |  |
| IIID | 3 (4%) | 2 (9%) | 1 (2%) | 0 (0%) |  |  |
| IVA | 9 (11%) | 3 (14%) | 6 (13%) | 0 (0%) |  |  |
| IVB | 12 (14%) | 6 (27%) | 3 (6%) | 3 (30%) |  |  |
| IVC | 24 (30%) | 6 (32%) | 14 (30%) | 4 (40%) |  |  |
| IVD | 22 (28%) | 2 (9%) | 20 (43%) | 0 (0%) |  |  |
| Metastatic disease sites |  |  |  |  |  |  |
| Brain | 22 (28%) | 2 (9%) | 20 (43%) | 0 (0%) | **0.0014** | 0.1 |
| Liver | 18 (23%) | 3 (14%) | 12 (26%) | 3 (30%) | 0.43 | 0.86 |
| Multi-site involvement<br>(≥3 organ sites) | 33 (42%) | 6 (27%) | 24 (51%) | 3 (30%) | 0.14 | 0.63 |
| Baseline LDH |  |  |  |  |  |  |
| Elevated (>upper limit of<br>normal) | 28 (35%) | 1 (5%) | 23 (49%) | 4 (40%) | **0.0018** | 0.7 |
| Normal | 35 (44%) | 16 (73%) | 15 (32%) | 4 (40%) |  |  |
| Not measured | 16 (20%) | 5 (23%) | 9 (19%) | 2 (20%) |  |  |
| ECOG score<sup>‡</sup> |  |  |  |  |  |  |
| 0 | 42 (53%) | 9 (41%) | 28 (60%) | 5 (50%) | 0.3 | 0.08 |
| 1 | 34 (43%) | 11 (50%) | 18 (38%) | 5 (50%) |  |  |
| 2 | 2 (3%) | 2 (9%) | 0 (0%) | 0 (0%) |  |  |
| Unknown | 2 (3%) | 0 (0%) | 1 (2%) | 0 (0%) |  |  |
| CMV serostatus |  |  |  |  |  |  |
| Positive | 51 (65%) | 14 (64%) | 30 (64%) | 7 (70%) | >0.99 | >0.99 |
| Prior therapy |  |  |  |  |  |  |
| Treatment naïve | 51 (65%) | 19 (86%) | 25 (53%) | 7 (70%) | **0.02** | 0.81 |
| Immunotherapy | 19 (24%) | 0 (0%) | 17 (36) | 2 (20%) | **0.0012** | 0.82 |
| Targeted therapy | 6 (8%) | 0 (0%) | 5 (11%) | 1 (10%) | 0.25 | 0.81 |
| Radiotherapy | 13 (59%) | 1 (5%) | 11 (23%) | 1 (10%) | 0.13 | 0.42 |
| Cytotoxic | 1 (1%) | 1 (5%) | 0 (0%) | 0 (0%) | NA | NA |

<sup>#</sup>Fisher's exact test, except as noted. <sup>^</sup>Mann-Whitney test. <sup>§</sup>Log-rank (Mantel-Cox) test (discrete data) or Cox Proportional Hazards Regression (continuous data).
<sup>‡</sup>RECIST, Response Evaluation Criteria in Solid Tumors; PR, partial response; CR, complete response; SD, stable disease; PD, progressive disease; CTCAE, Common Terminology Criteria for Adverse Events version 5.0 (45); AJCC, American Joint Committee on Cancer (44); ECOG, Eastern Cooperative Oncology Group (77); LDH, Lactate dehydrogenase; CMV, cytomegalovirus.

Given recent findings on the association between CMV seropositivity and improved ICB outcomes (24), we assessed this feature in our cohort. 65% of patients (51/79) were seropositive for CMV as defined by the presence of anti-CMV capsid IgG by ELISA (**Supplementary Fig. 5B**). We observed no differences in clinical features (**Supplementary Table 8)** or outcomes including PFS and OS between CMV^+^ and CMV^-^ patients (**Fig. 1C, F Supplementary Fig. 5C-E**), although a ‘crossover’ OS curve precluded robust statistical testing (**Supplementary Fig. 1D**). There remained no statistically significant association between CMV serostatus and OS when assessed within treatment cohorts, although we note that cohort numbers for nivo/pembro (n=22) and rela+nivo (n=10) may have impacted this finding (**Supplementary Fig. 5I**).

To assess the impact of age on clinical and immunological response to ICB, the cohort was split into younger (<65 years age) and older (≥65 years) patients. The younger cohort (n=30) had a median age of 55 (range 25-63) and the older cohort (n=49) had a median age of 77 (66–92). Older patients more often received nivo/pembro and rela+nivo (*p*=0.018) and had a worse ECOG performance score (*p*=0.039) than younger patients, but other clinical features were not significantly different (**Supplementary Table 9**). Furthermore, there were no differences in clinical outcomes between age groups (**Fig. 1C, G; Supplementary Fig. 5C, F-H**), and neither age nor CMV status associated with OS, PFS, or TFS in Cox proportional hazards tests incorporating ECOG, treatment, age, and CMV serostatus (**Fig. 1E-F, Supplementary Fig. 5A**).

### CMV is associated with expanded functional effector memory but stable naive CD8+ T cell numbers

Given that CMV is known to cause significant alterations to the T cell compartment (15,18) and T cells are key ICB effectors, we sought to validate whether CMV seropositivity affected pre-treatment and treatment-emergent immune phenotype in patients with advanced melanoma. Using a modified TruCount flow cytometry panel performed on fresh whole blood, we first asked whether the absolute abundance of major innate and adaptive immune subsets differed between patients with and without CMV infection. We observed higher circulating lymphocyte counts in CMV^+^ versus CMV^-^ patients (median 1411 vs. 1112 cells/μL, respectively, *p*=0.0030) (**Fig. 2A**), resulting from higher T-cell counts (1075 cells/μL vs. 764 cells/μL, respectively, *p*=0.0025; **Fig. 2B**). This resulted in a decreased neutrophil-to-lymphocyte ratio (NLR) and an increased lymphocyte-to-monocyte ratio (LMR) in CMV^+^ patients, a phenotype previously associated with longer OS following ICB (54,55) (**Supplementary Fig. 6A**). There were otherwise no significant differences in the abundance of total leukocytes, granulocytes (neutrophils and eosinophils), monocytes (classical, intermediate, and non-classical subsets), B, or NK cells between CMV^+^ and CMV^-^ patients (**Fig. 2B, Supplementary Fig. 6B-C**).

**Figure 2.**
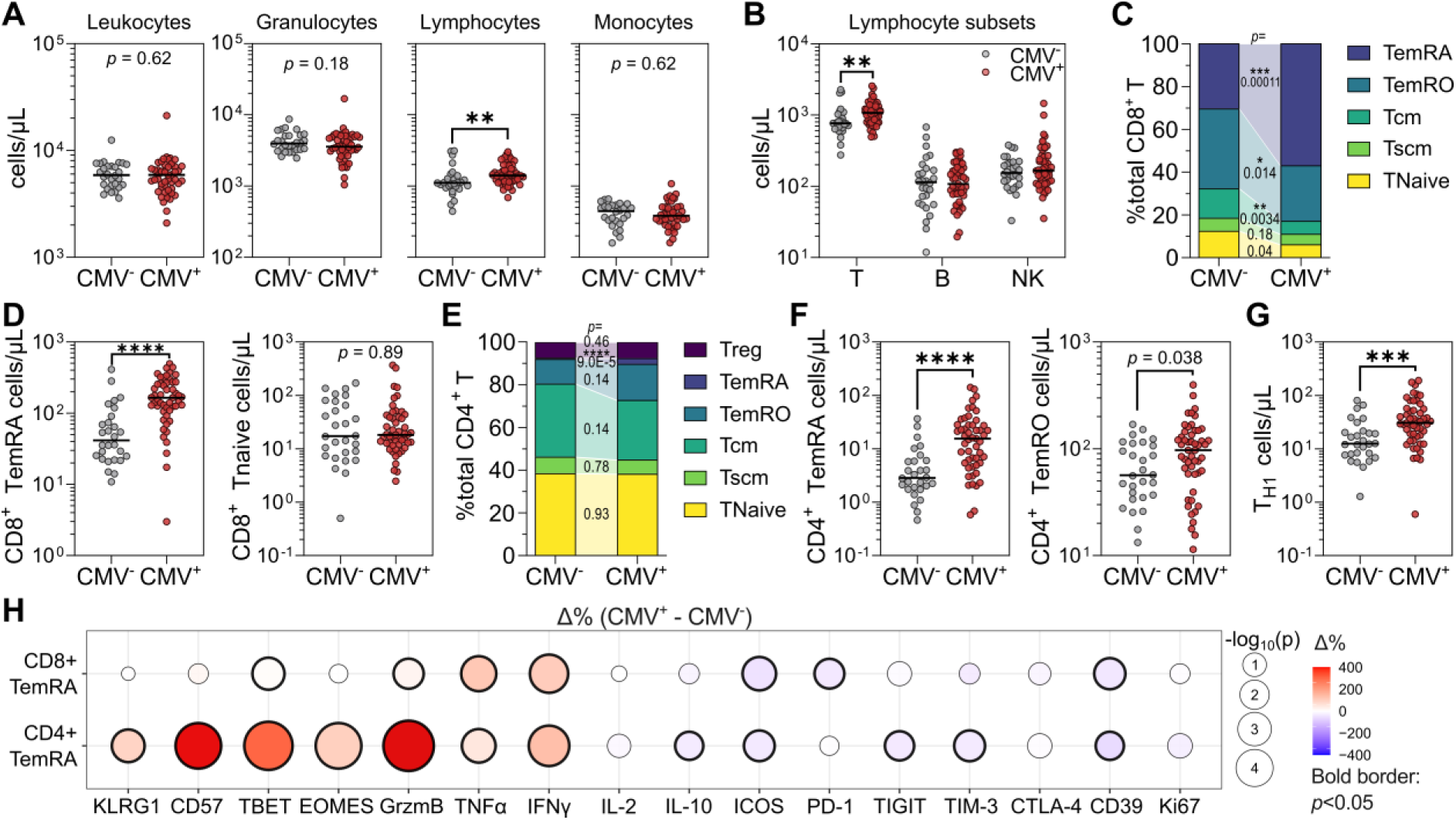
Pre-treatment immune phenotype of CMV^+^ and CMV^-^ melanoma patients. (**A**) Absolute counts per µL of blood of leukocytes, granulocytes, lymphocytes, and monocytes in CMV^-^ (n=28) and CMV^+^ (n=51) patients before treatment. (**B**) Absolute counts of T, B, and NK cells. (**C**) Proportional abundance of CD8^+^ T subsets. Mann-Whitney *p* values are displayed in joining segments. (**D**) Absolute abundance of CD8^+^ TemRA and Tnaive cells. (**E**) Proportional CD4^+^ T subset abundance as per (C). (**F**) Absolute abundance of CD4^+^ TemRA, TemRO and (**G**) T_H1_ cells. (**H**) CMV-related differences in proportional marker expression by CD8^+^ and CD4^+^ TemRA. Balloon size represents -log_10_(*p*) value, with *p*-values < 0.05 marked by a bold border. Colour represents percent delta expression (CMV^+^ - CMV^-^); positive (red) values denote features upregulated in CMV^+^ patients. All plots display median value, *p* values from Mann-Whitney tests.

To assess differences in T-cell phenotypes, we performed high-parameter spectral flow cytometry on paired samples from all 79 patients using two previously developed panels (46). This approach allowed assessment of absolute cell counts per μL of whole blood and of the proportional abundance of 12 phenotyping markers and 7 cytokines across 17 distinct T-cell subsets, including γδT and αβT cells and subsets therein: CD4^+^ and CD8^+^ T cells; CD4^+^ T regulatory (Treg) and follicular helper (T_FH_), and CD4^+^ and CD8^+^ naive T (Tnaive; *TN*), stem cell-like memory (Tscm; *TS_B_M_p_*), central memory (Tcm; *TS_B_M*), effector memory RO (TemRO; *TD_B_M*), and effector memory cells re-expressing CD45RA (TemRA; *CD45RA^+^ TD_B_M*) (**Supplementary Tables 4-5**). Italicized text denotes T cell subset names as per the recently introduced T-cell nomenclature guidelines (56); this text will henceforth use the previous nomenclature for clarity. We also enumerated T_H1_ (IFNγ^+^ IL-17A^-^), T_H2_ (IFNγ^-^ IL-17A^-^ IL-4^+^), and T_H17_ (IFNγ^-^IL-17A^+^) within CD4^+^ T cells.

CMV^+^ patients had higher total CD8^+^ T-cell numbers than CMV^-^ patients, but similar numbers of CD4^+^ T cells, resulting in a significantly lower CD4:CD8 T cell ratio (**Supplementary Fig. 6D-E**). As reported in healthy individuals, CMV^+^ patients had proportional and absolute expansions of CD8^+^ TemRA (median CMV^+^ 165 cells/μL, 51% of total CD8^+^ T; median CMV^-^ 41 cells/μL, 27%, *p*=2.8 x 10^-6^ and 0.00011, respectively) (**Fig. 2C**). Proportional CD8^+^ TemRA abundance correlated with CMV IgG titer (**Supplementary Fig. 6F**). The proportional abundance of CD8^+^ Tnaive cells was correspondingly lower in CMV^+^ patients (7% vs. 15% in CMV^-^ patients, *p*=0.04), despite similar absolute abundances (median CMV^+^ 18 cells/μL vs. CMV^-^ 17 cells/μL, *p*=0.89) (**Fig. 2C-D**). Similarly, while the proportional abundances of CD8^+^ Tcm (*p*=0.0034) and TemRO (*p*=0.014) cells were also lower in CMV^+^ patients, absolute abundances were similar. These differences in CMV^+^ patients also resulted in higher ratios of CD8^+^ T cells to Treg (**Supplementary Fig. 6G**).

The majority of CD8^+^ TemRA from both CMV^+^ and CMV^-^ patients expressed CD57 (68% and 57%, respectively; *p*=0.30) and KLRG1 (86% and 82%, *p*=0.87), but did not express Ki67 (1.98% and 1.85%, *p*=0.31), suggesting that these cells were mostly non-proliferative. However, CD8^+^ TemRA retained high levels of effector molecules, most of which were expressed more frequently in CMV^+^ patients: IFNγ (*p*=0.00019), TNFα (*p*=0.0014), and granzyme B (*p*=0.018) (**Fig. 2G**). CMV^+^ patients hence had an increased frequency of polyfunctional CD8^+^ T cells co-expressing 2 or more of the markers IFNγ, TNFα, granzyme B, and IL-2 (**Supplementary Fig. 6H**).

Furthermore, CMV^+^ patients had a higher frequency of TBET expression (*p*=0.0068), lower frequency of PD-1 (35% vs. 22%, *p*=0.020), and trends for lower TIGIT and TIM-3 expression (**Fig. 2H**), demonstrating that the higher numbers of CD8^+^ TemRA in CMV^+^ patients represented functional, non-exhausted CD8^+^ T cells. Notably, 48% of proliferative Ki67^+^ CD8^+^ T cells expressed PD-1 in CMV^+^ patients as opposed to 60% in CMV^-^ patients (*p*=7.8 x 10^-5^; **Supplementary Fig. 6I**), indicating that proliferative cells in CMV^+^ patients may be less functionally exhausted. Alongside changes to CD8^+^ TemRA, CD8^+^ TemRO also had a higher frequency of IFNγ (*p*=0.0008) and a lower frequency of PD-1 expression (*p*=0.0032). Hence, CMV^+^ patients carried more highly functional effector CD8^+^ T cells.

### CMV^+^ patients have more terminally differentiated cytotoxic CD4^+^ T cells

Alterations to CD4^+^ T-cell subsets were also noted in CMV^+^ individuals (**Fig. 2E**), including 5.4-fold higher CD4^+^ TemRA numbers (16 cells/μL vs 3 cells/μL in CMV^-^ patients; *p*=1.5 x 10^-5^) and nearly 2-fold higher CD4^+^ TemRO numbers (97 cells/μL vs 56 cells/μL in CMV^-^; *p*=0.038) (**Fig. 2F**). Furthermore, T_H1_, but not T_H2_ or T_H17_, were higher in CMV^+^ patients (*p*=0.0007). This may be driven by TBET expression, which was significantly higher in total CD4^+^ T cells (median 3% vs 9%, *p*=1.4 x 10^-5^), but especially so in TemRO and TemRA (2.6 and 4.0-fold higher, *p*=5.6 x 10^-5^ and 7.0 x 10^-8^, respectively) (**Fig. 2G, Supplementary Fig. 6J**). The numbers of the remaining CD4^+^ T cell subsets, Treg, T_FH_, and γδT cells were not different between CMV^+^ and CMV^-^ patients.

The expanded CD4^+^ TemRA population in CMV^+^ patients also demonstrated markedly higher expression of effector cytokines including granzyme B (16.5% vs 1.6%, *p*=9.2 x 10^-9^), IFNγ (15.4% vs 6.6%, *p*=4.1 x 10^-5^), and TNFα (14% vs 9.2%, *p*=0.003), as well as more frequent expression of KLRG1, CD57, TBET, and EOMES, and less prevalent expression of ICOS, TIGIT, TIM-3, and CD39 (**Fig. 2H**). Similar alterations were noted for CD4^+^ TemRO (**Supplementary Fig. 6K**). Hence, CMV infection in melanoma patients was associated with higher numbers of functional CD4^+^ and CD8^+^ cells with limited proliferative but enhanced effector potential, and did not affect levels of circulating naive T cells.

### Older patients have lower B-cell numbers with signs of immune ageing

Age is also known to affect immune cell phenotype (4), so we sought to validate whether an immune ageing signature was present in older patients with advanced melanoma. We found no significant age-based differences in the numbers of circulating leukocytes (median 5529 and 5934 cells/µL for young and old, respectively, *p*=0.78), granulocytes (3632 and 3885 cells/µL, *p*=0.56), total lymphocytes (1364 and 1249 cells/µL, *p*=0.50), or monocytes (349 and 417 cells/µL, *p*=0.17) (**Fig. 3A**), or major monocyte and granulocyte subsets (**Supplementary Fig. 7A-B**). However, older patients had a significantly lower lymphocyte-to-monocyte ratio (LMR; 3.1 vs 4.4 in young, *p*=0.0038, **Fig. 3B**), and this decrease correlated with increasing age (Spearman r=-0.26, *p*=0.027, **Fig. 3C**).

**Figure 3.**
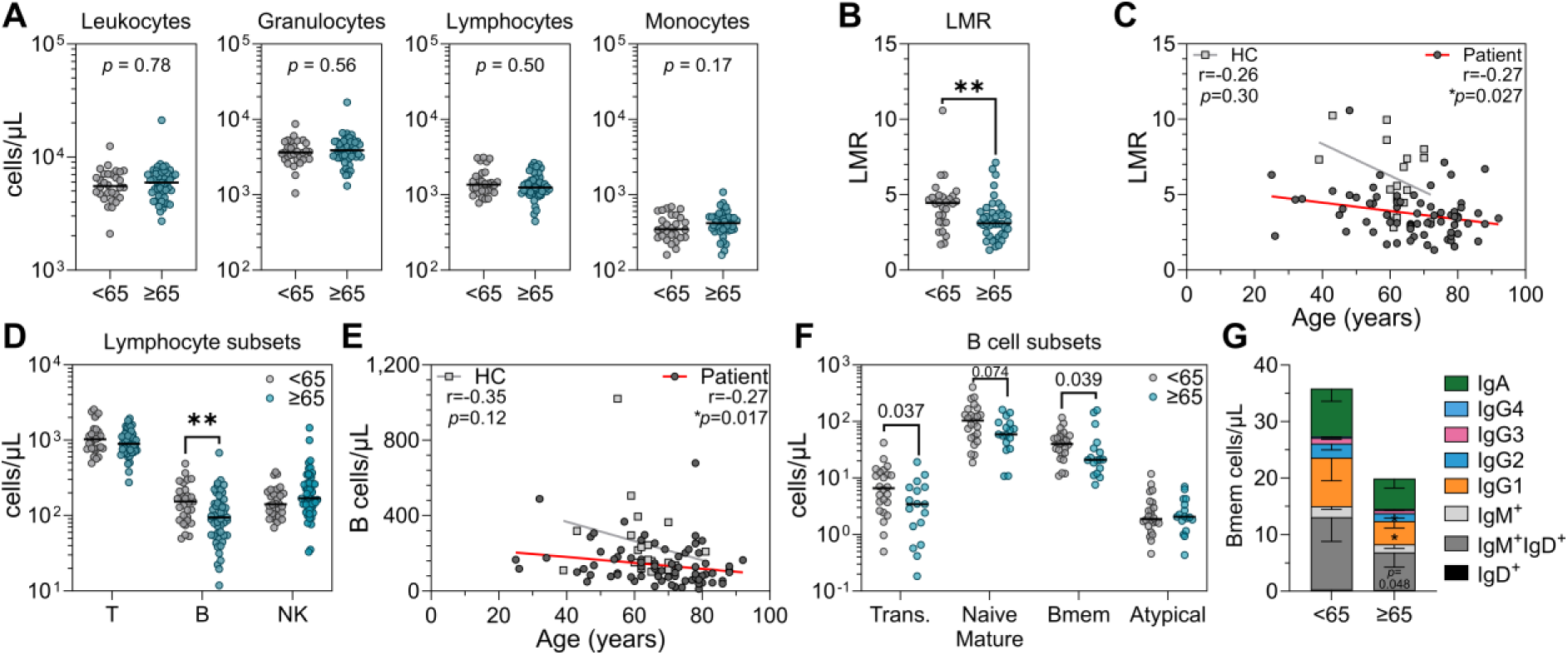
Pre-treatment immune phenotype in melanoma patients stratified by age. (**A**) Absolute counts per µL of blood of leukocytes, granulocytes, lymphocytes, and monocytes in younger (<65 years old, n=30) and older (≥65, n=49) patients before treatment. (**B**) Lymphocyte to monocyte ratio (LMR). (**C**) Spearman correlation of LMR and age in patients (n=79) and healthy controls (HC; n=21). (**D**) Absolute counts of T, B, and NK cells. (**E**) Spearman correlation between B cell counts and age in patients (n=79) and healthy controls (HC; n=21). (**F**) Absolute counts of transitional (trans.), naive mature, memory (Bmem), and CD21^-^ CD11c^+^ atypical B cells. (**G**) Absolute abundance of Ig subclasses. Plots represent median, *p* values from Mann-Whitney tests unless otherwise noted.

Of the major lymphocyte subsets, only B-cell numbers were significantly lower in elderly patients (*p*=0.0093, **Fig. 3D**), again correlating inversely with age (r=-0.27, *p*=0.017, **Fig. 3E**), whereas T-cell abundance did not (**Supplementary Fig. 7C**). To assess alterations to the B-cell phenotype, we performed high-parameter spectral flow cytometry on available paired pre- and on-treatment samples from 41 patients receiving ipi+nivo, as well as single samples from healthy controls (n=16) for descriptive comparison. This panel allowed us to enumerate total CD19^+^ B cells; transitional B cells, plasmablasts, naive mature and memory (Bmem) B cells; CD21^-^ CD11c^+^ atypical B cells (atBC), and the proportions of Bmem expressing various Ig isotypes and IgG subclasses (**Supplementary Fig. 4**). We also included the markers TIM-1 and PD-L1 for their relevance to ICB response (57,58), as well as fluorescently-labelled tetramers of the influenza HA protein to assess the effect of ICB on anti-viral immune memory.

Within the cohort of 41 patients receiving ipi+nivo, we again observed lower total B-cell counts in aged individuals (median 88 cells/μL vs. 152 cells/μL, *p*=0.055, **Supplementary Fig. 7D**), including transitional (*p*=0.037), naive mature (*p*=0.074), and Bmem (*p*=0.039) subsets (**Fig. 3F**). In contrast, the frequencies of atBCs within Bmem tended to be higher in older patients (median 7.2% vs. 4.7%, *p*=0.07) due to a stable absolute abundance with age (**Fig. 3F**). Similarly, the influenza-specific B-cell population of elderly patients contained a higher proportion of atBCs (median 37% vs. 19%, *p*=0.061), but the absolute cell numbers were not different, and total atBC abundance did not correlate with age (**Supplementary Fig. 7E-F**). Regardless of age, atBCs were enriched for expression of PD-L1 as compared to conventional Bmem (*p*<0.0001, **Supplementary Fig. 7G**). The reduced Bmem numbers in older patients encompassed all Ig isotypes and IgG subclasses (**Fig. 3G**), of which the IgG1^+^ (*p*=0.027), and IgG2^+^ populations (*p*=0.018) were most significantly reduced. Overall, we observed evidence of minor age-related differences in the circulating B cell compartment.

### Lower CD8^+^ naive T cells and moderately higher TemRA in older patients

As T cells are the principal targets of ICB and are known to be impacted by immune ageing (4), we compared T-cell phenotype between younger and older patients. While there were no changes in the abundance of total CD4^+^ or CD8^+^ T cells when assessing pre-treatment samples from all 79 patients, older patients had a significant proportional shift from naive to TemRA cells in their CD8^+^ T cell compartment. 4.2% of total CD8^+^ T cells were naive in older patients as opposed to 17.2% in younger patients (*p*=2.5 x 10^-5^), while TemRA cells expanded from 26.1% to 51.0% in older patients (*p*=1.8 x 10^-5^) (**Fig. 4A**). This shift was due to both an absolute decrease in naive cells (52 to 13 cells/µL in young and old patients, respectively, *p*= 6.9 x 10^-8^) and an increase in TemRA cells (106 to 140 cells/µL, *p*=0.032) (**Fig. 4B**). These changes correlated significantly with age (naive spearman r=-0.62, *p*<0.0001; TemRA r=0.23, *p*=0.043) (**Supplementary Fig. 7H-I**). CD8^+^ TemRA cell expansion resulted in older patients having a greater ratio of PD-1^+^ CD8^+^ effector memory cells to PD-1^+^ Treg cells (6.6 versus 4.4, *p*=0.015), a feature previously reported to associate with improved response to ICB (59) (**Fig. 4D**). A decreased abundance of CD8^+^ Tcm in older patients was also observed (30 versus 14 cells/μL, *p*=0.0054).

**Figure 4.**
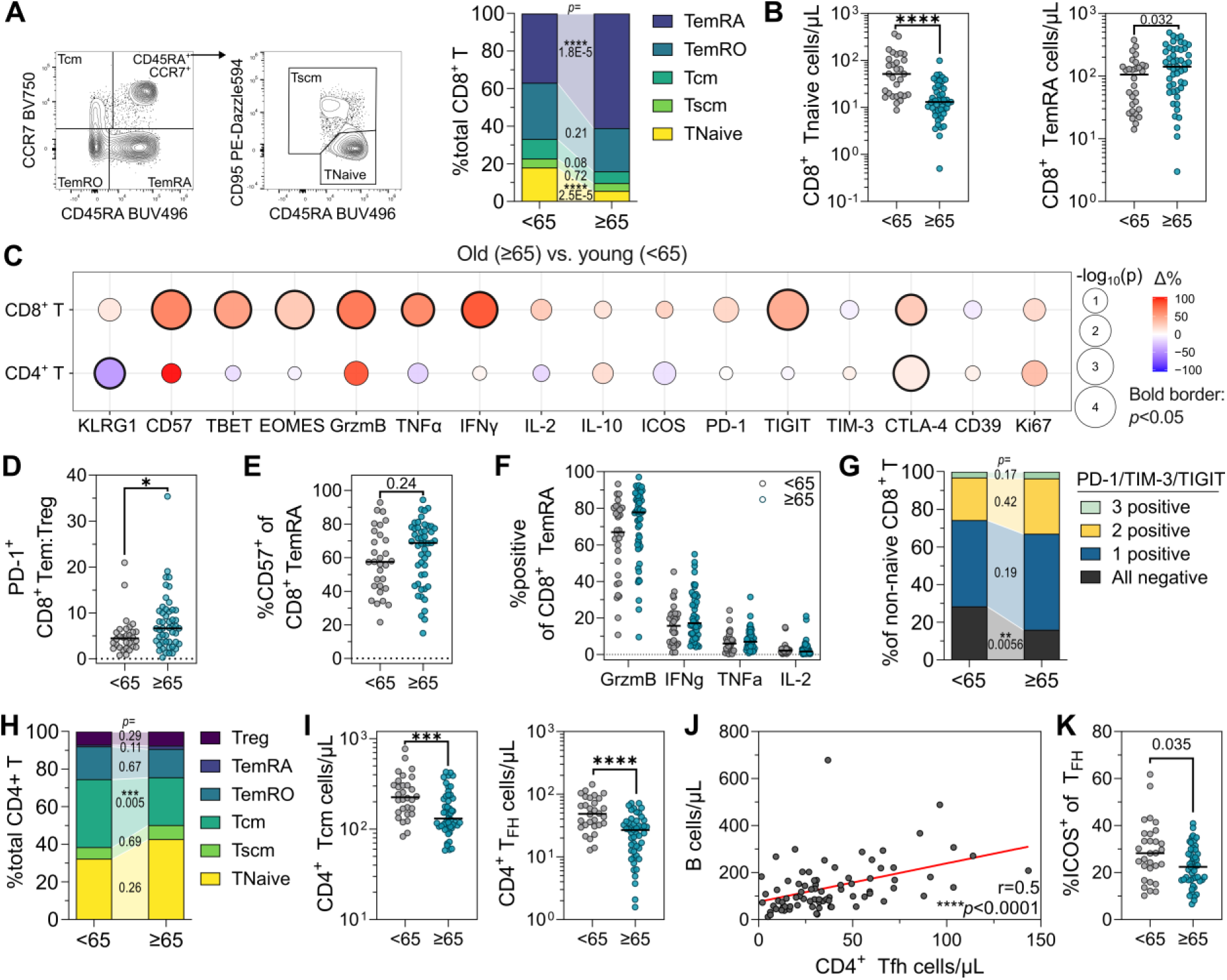
Pre-treatment T-cell phenotype of patients stratified by age. (**A**) Representative gating and proportional abundance of CD8^+^ T in older (≥65 years old, n=49) compared to younger (<65, n=30) patients. Mann-Whitney *p* values are displayed in joining segments. (**B**) Absolute abundance of CD8^+^ Tnaive and TemRA cells. (**C**) Age-related differences in proportional marker expression by total CD8^+^ and CD4^+^ T cells. Balloon size represents - log_10_(*p*) value, with *p*-values < 0.05 marked by a bold border. Colour represents percent delta expression (old - young); positive values denote features upregulated in older patients. (**D**) Ratio of PD-1^+^ CD8^+^ T effector memory cells over PD-1^+^ Treg. (**E**) CD8^+^ TemRA expression of the senescence marker CD57 and (**F**) cytokines. (**G**) Boolean co-expression of the inhibitory molecules PD-1, TIM-3, and TIGIT. (**H**) Proportional abundance of CD4^+^ T subsets. Mann-Whitney *p* values are displayed in joining segments. (**I**) Absolute abundance of CD4^+^ Tcm and T_FH_ cells. (**J**) Spearman correlation of T_FH_ and B cell abundance in patients (n=79). (**K**) Percent expression of the co-stimulatory marker ICOS by T_FH_ cells. All plots display median value, *p* values from Mann-Whitney tests unless otherwise noted.

Total CD8^+^ T cells of older patients had higher proportions expressing IFNγ (84% increase, *p*=0.0042), granzyme B (70% increase, *p*=0.0018), TNFα (62% increase, *p*=0.017), CD57 (64% increase, *p*=0.0009), TIGIT (45% increase, *p*=0.0004), EOMES (30% increase, *p*=0.0014), and TBET (51% increase, *p*=0.0029) than younger patients (**Fig. 4C**). However, these changes predominantly reflected the overall shift from naive to terminal effector memory cells. The only phenotypic changes within the expanded TemRO and TemRA subsets were increased frequencies of TIGIT (51 to 65%, *p*=0.0041) and CTLA-4 (24 to 31%, *p*=0.0015), and decreased KLRG1 (80 to 63%, *p*=0.0069) in aged CD8^+^ TemRO (**Fig. 4E-F, Supplementary Fig. 7J**). The proportions of total CD8^+^ T cells expressing CTLA-4 and PD-1 also trended higher (28% increase, *p*=0.034; 24% increase, *p*=0.096, for CTLA-4 and PD-1 respectively) in older patients (**Fig. 4C**). Boolean co-expression analysis of PD-1, TIM-3, and TIGIT showed that more non-naive CD8^+^ T cells from older patients expressed one or more of these inhibitory markers (*p*=0.0058), potentially indicating a moderate increase in exhaustion levels with age (**Fig. 4G**).

Given that CMV infection also caused significant changes to the CD8^+^ compartment, we performed multiple linear regression to independently assess the role of ageing and CMV infection on CD8^+^ naive and TemRA abundance. This demonstrated that CD8^+^ naive T cell abundance decreased by 2.15 cells/μL per year of age (95% CI 1.30 – 3.00, *p*<0.0001) but was not affected by CMV status (*p*=0.60). Conversely, CD8^+^ TemRA count was increased by both CMV seropositivity (106 cells/μL, 95% CI 56-156, *p*<0.0001) and age (1.68 cells/μL per year, 95% CI 0.038-3.31, *p*=0.045). Hence, while CMV both expanded TemRA cells and increased their effector functionality, age depleted naive cells and led to moderate TemRA cell expansion without affecting phenotype.

### Decreased T_FH_ cell abundance in older patients

CD4^+^ T cells were differently affected by ageing. No decrease in naive CD4^+^ T cells was observed; rather, there was a proportional (35 to 28%, *p*=0.0053) and absolute (225 to 131 cells/μL, *p*=0.0009) decrease in CD4^+^ Tcm (**Fig. 4H-I**). Significantly lower numbers of CD4^+^ T_FH_ (49 versus 27 cells/μL, *p*=1.4 x 10^-5^) and T_H17_ (11 versus 6 cells/μL, *p*=0.0054) were also observed in older patients (**Fig. 4I, Supplementary Fig. 7K**). The declining abundance of T_FH_ and B cells was strongly correlated (Spearman r=0.5, *p*<0.0001, **Fig. 4J**), and T_FH_ from aged patients had decreased ICOS expression (*p*=0.035, **Fig. 4K**), potentially indicating that T cell defects drive impaired humoral immunity in older patients. There were fewer changes to CD4^+^ marker expression, with only CTLA-4 increasing 11% (*p*=0.0049) attributable to Tnaive, Tscm, TemRO, and TemRA subsets, and KLRG1 decreasing 44% (*p*=0.032) attributable to Tscm, Tcm, and TemRO cells (**Fig. 4C**).

### Ipi+nivo induced greater immune changes than PD-1 ± LAG-3 blockade

Despite characterizing significant age- and CMV-associated effects on immune composition in our cohort of patients with advanced melanoma prior to ICB treatment, clinical outcomes to ICB were notably similar across age and CMV groups. Therefore, we next sought to determine whether CMV or ageing affected the immunological changes induced by ICB. Given that the effects of different ICB treatments are at least partially distinct (60–63), we first defined pre- to on-treatment immune changes between patients who had received either one cycle of nivo/pembro (n=22), ipi+nivo (n=47), or rela+nivo (n=10), agnostic of age and CMV serostatus. Principal component analysis (PCA) showed no distinct separation of immune phenotype between patient groups before beginning treatment. However, it was evident that the on-treatment phenotype of patients receiving ipi+nivo clustered distinctly from those receiving nivo/pembro and rela+nivo, implying that the addition of CTLA-4 blockade induced immune changes distinct from or of greater magnitude than PD-1 ± LAG-3 blockade (**Fig. 5A**). We quantified this by calculating Bray-Curtis dissimilarity and utilizing PERMANOVA and PERMDISP to assess immune composition variance. Treatment group explained 5.3% of immune composition variation before treatment (pseudo-F = 2.12, R^2^ = 0.053, *p*=0.0008), but in support of PCA data, this effect size increased on-treatment to 10.0% (pseudo-F = 4.23, R^2^ = 0.10, *p*=0.0001), suggesting that there was greater divergence in immune composition between treatment groups on-treatment. Importantly, there was no significant difference in dispersion pre-(*p*=0.14) or on-treatment (*p*=0.63), supporting the validity of these comparisons.

**Figure 5.**
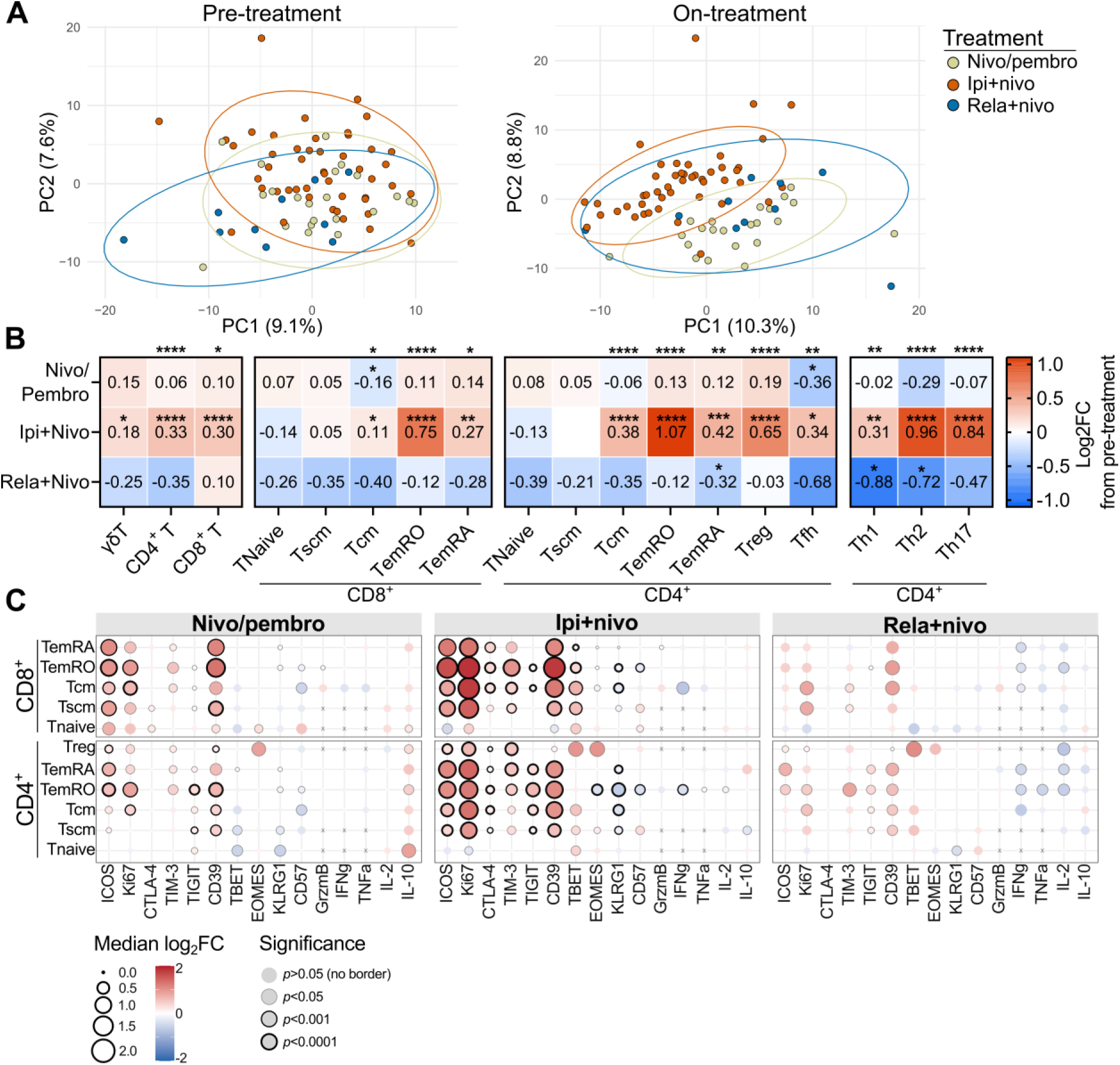
Divergent immunological effects of PD-1 mono- and combination therapies. (**A**) Pre- and on-treatment principal component analysis (PCA) plots coloured by treatment type; ipi+nivo (n=47), nivo/pembro (n=22), and rela+nivo (n=10). PCA plots use proportional data. (**B**) Pre- to on-treatment log_2_ fold change in the absolute abundance of T cell subsets stratified by treatment. Positive (red) values represent populations expanded on-treatment. *P* values from paired Wilcoxon tests; \**p*<0.0167, \*\**p*<0.01, \*\*\**p*<0.001, \*\*\*\**p*<0.0001. (**C**) Pre- to on-treatment log_2_ fold change in proportional marker expression by CD8^+^ and CD4^+^ T subsets stratified by treatment. Balloon size and colour represent log_2_FC value, while *p* values are denoted by balloon border. All data denotes median. *P* values from paired Wilcoxon tests.

To explore which features contributed to this divergence, we first examined changes to the absolute abundance of immune subsets. Ipi+nivo treatment resulted in significant expansions of both CD4^+^ and CD8^+^ T cells, most notably CD4^+^ TemRO (median Log_2_FC=1.07, *p*=2.9 x 10^-12^) and CD8^+^ TemRO (Log_2_FC=0.75, *p*=1.0 x 10^-8^), as well as both CD4^+^ and CD8^+^ Tcm and TemRA, and Treg (**Fig. 5B**). The expansion of CD4^+^ and CD8^+^ subsets correlated strongly with their pre-treatment expression of PD-1, but only in patients treated with ipi+nivo (r=0.82, *p*=0.0033), implying that PD-1 expression potentiates CTLA-4-driven increases in abundance (**Supplementary Fig. 8A**). While nivo/pembro induced a moderate and non-significant T-cell expansion, rela+nivo generally resulted in a slight reduction in T-cell abundance, including significant decreases in T_H1_ (*p*=0.014) and T_H2_ (*p*=0.0059) cells. Conversely, ipi+nivo led to significant increases in T_H1_, T_H2_, and T_H17_ cells (*p*=0.0031, 1.1 x 10^-6^, and 6.8 x 10^-7^, respectively) (**Fig. 5B**). Ipi+nivo also resulted in an expansion of eosinophils (Log_2_ fold change = 1.09, *p*=2.4 x 10^-5^) and classical and intermediate monocytes (Log_2_FC = 0.29 and 0.72, *p*=2.6 x 10^-5^ and 0.0002, respectively), and a decrease in B cells (Log_2_FC=-0.26, *p*=0.037). These changes were not observed, or were not significant, in patients receiving nivo/pembro or rela+nivo (**Supplementary Fig. 8B-D**). Hence, PD-1 and CTLA-4 blockade induced changes of greater magnitude and affecting more subsets than PD-1 blockade alone or in combination with LAG-3 blockade.

We next assessed pre- to on-treatment Log_2_ fold change in the expression frequency of non-subset defining markers (see *x* axis of **Fig. 5C**) across CD8^+^ and CD4^+^ T subsets to determine if the phenotype or effector functionality of subsets differed on-treatment. There were 27, 56, and 14 upregulated markers (*p*<0.05, Log_2_FC>0.5) following one cycle of nivo/pembro, ipi+nivo, and rela+nivo, respectively, and 5, 4, and 5 features downregulated (*p*<0.05, Log_2_FC<-0.5), respectively, although we note that the size of the rela+nivo cohort likely limited our power to detect differences (**Supplementary Table 10**). Of the 31 additional markers upregulated in patients receiving ipi+nivo compared to patients receiving nivo/pembro, 20 pertained to CD4^+^ T, 4 to CD8^+^ T, 5 to γδT, and 2 to B cells. This supports previous findings suggesting that CTLA-4 blockade predominantly affects CD4^+^ T cells (64), a paradigm that may be explained by the higher portion of stimulated CD4^+^ T cells expressing CTLA-4 than CD8^+^ T cells (**Supplementary Fig. 8E**).

All treatments generally increased the expression frequency of ICOS, Ki67, and CD39 across multiple CD4^+^ and CD8^+^ subsets, but these changes were typically of larger magnitude and more significant in patients treated with ipi+nivo (**Fig. 5C**). CD8^+^ TemRO typically demonstrated the largest upregulation of these markers followed by CD4^+^ and CD8^+^ Tcm and TemRA, reflecting that the largest absolute expansions were also observed for these subsets (**Fig. 5B**), and that these subsets expressed the highest levels of PD-1 prior to ICB treatment (**Supplementary Fig. 8E**). Compared to nivo/pembro, ipi+nivo additionally resulted in greater upregulation of CTLA-4, TBET, TIGIT, and TIM-3 across multiple CD4^+^ and CD8^+^ subsets. Downregulated features following ipi+nivo included IFNγ and KLRG1 expression by Tcm and TemRO cells, while rela+nivo tended to downregulate effector cytokines IFNγ, TNFα, and IL-2 on CD4^+^ TemRO and TemRA cells (**Fig. 5C**). Hence, it seems that PD-1 blockade upregulated a shared set of markers including Ki67, CD39, and ICOS, and to a lesser extent EOMES and TIM-3, while CTLA-4 addition induced changes to more features and of greater magnitude than LAG-3 addition.

### Ipi+nivo decreases B cell abundance and proportionally enriches for atypical B cells

As ipi+nivo treatment decreased absolute B cell count (**Supplementary Fig. 9A**), we also assessed changes to B cell phenotype within this cohort. Total B cells decreased from 128 to 112 cells/μL on-treatment (*p*=0.0056), resulting predominantly from decreases to circulating naive mature (*p*=0.0007) and memory (*p*=0.0033) B cells (**Supplementary Fig. 9B**). Within Bmem, the abundance of all antibody isotypes decreased with the exception of IgM (**Supplementary Fig. 9C**). While the abundance of conventional B cells was reduced on-treatment (*p*=0.0015) (**Supplementary Fig. 9D**), atypical B cell numbers remained stable on-treatment and so were proportionally enriched (*p*=3.2 x 10^-8^; **Supplementary Fig. 9E**). We observed no difference in ICB-induced B cell alterations by age or CMV serostatus.

### Neither age nor CMV markedly alter the effect of ipi+nivo on peripheral immunity

As clinical response did not differ by CMV serostatus or age, we hypothesized that immunological responses to ICB would be independent of CMV status or age. Given that ipi+nivo induced the most significant immune changes and was the largest cohort, we focused on this group of patients and observed no difference in on-treatment T cell expansion between patients split by both age and CMV status (**Fig. 6A**), nor did this expansion correlate with age (Spearman r=0.19, *p*=0.20; **Fig. 6B**). There was a trend for older patients to experience greater CD8^+^ T cell expansion on treatment (Log_2_FC 0.51 vs. 0.16, *p*=0.046), driven by expanding CD8^+^ Tcm (0.48 vs 0.013, *p*=0.035) (**Fig. 6C**). However, ipi+nivo induced equivalent CD8^+^ TemRA cell expansion in younger and older patients, and so this subset remained enriched in older patients on-treatment. CD4^+^ T cell expansion and CD4:CD8 T cell ratio (*p*=0.12) were unaffected by age (**Fig. 6D**). Neither CD8^+^ nor CD4^+^ T cell expansion was different between CMV^-^ and CMV^+^ patients (**Fig. 6E-F**). Furthermore, treatment-induced changes to the expression of activation, proliferation, inhibition, differentiation, and cytokine markers were largely equivalent between young and old patients, with the exception of ICOS and granzyme B (**Fig. 6G**). Younger patients upregulated ICOS on CD8^+^ (*p*=0.0077) and CD4^+^ (*p*=0.0023) TemRO cells to a greater degree, whereas older patients tended toward greater upregulation of granzyme B on CD8^+^ TemRO (*p*=0.021) and total CD4^+^ T cells (*p*=0.024; **Fig. 6H-I**). CMV serostatus similarly had minimal effect on treatment-induced changes, other than marginally lower upregulation of TBET (*p*=0.032) and TNFα (*p*=0.022) frequency within CD4^+^ T cells from CMV^+^ patients (**Fig. 6J-K; Supplementary Fig. 8F**). There was also no age- or CMV-related difference in Ki67 upregulation within CD4^+^ Tcm or TIM-3 expression by CD8^+^ T cells, two previously reported features associated with response and toxicity to ipi+nivo ICB (**Supplementary Fig. 8G-H**) (43). Hence, individuals treated with ipi+nivo experienced highly similar treatment-induced immune changes regardless of CMV status or age, effectively preserving any pre-treatment age- and CMV-related differences in T cell profiles.

**Figure 6.**
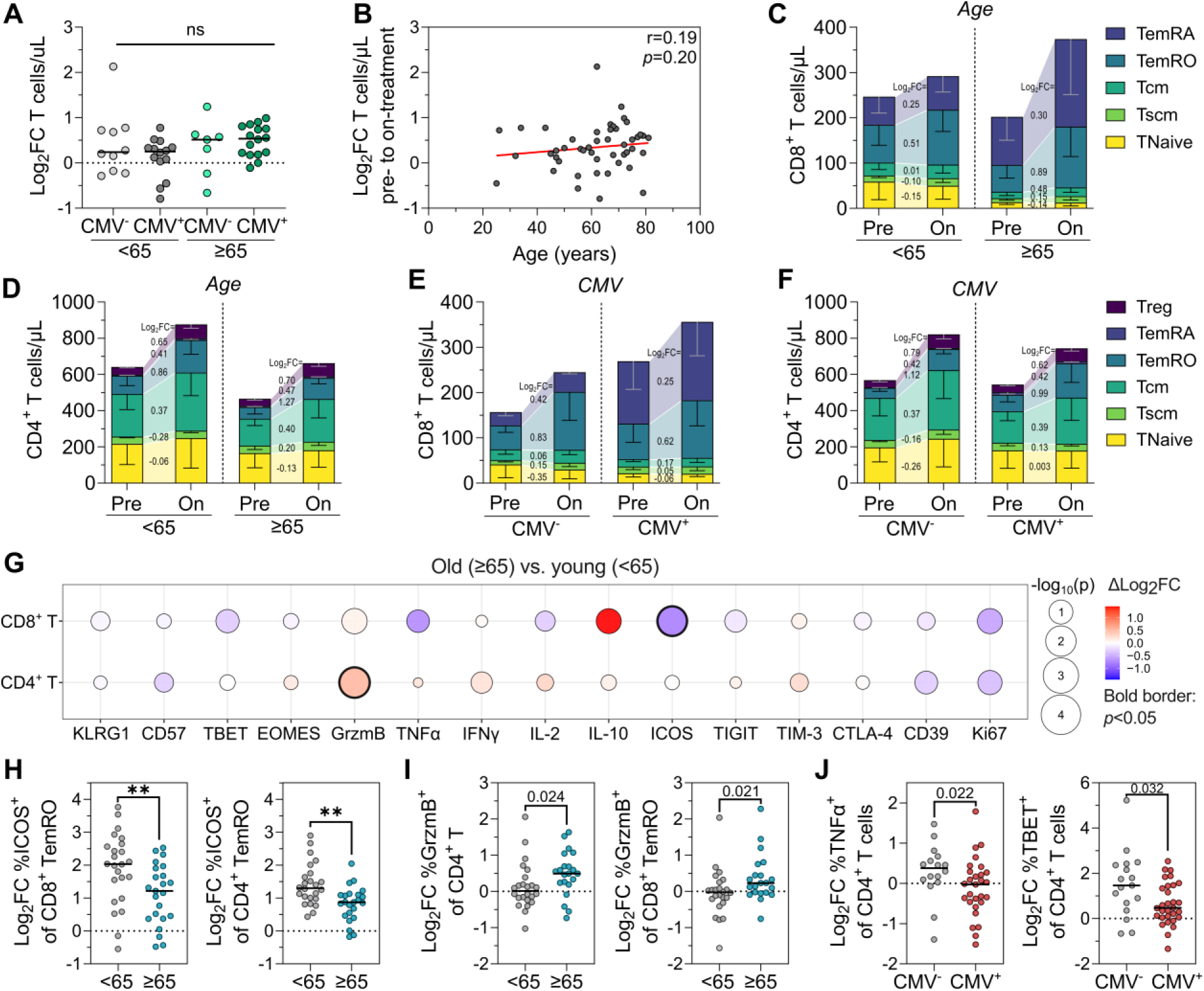
Age and CMV status have no effect on immune changes induced by ipi+nivo. (**A**) Pre- to on-treatment log_2_ fold change (Log_2_FC) in absolute T cell abundance between young CMV^-^ (n=10), young CMV^+^ (n=14), old CMV^-^ (n=7), and old CMV^+^ (n=16) patients receiving ipi+nivo (Kruskal-Wallis *p*=0.22). (**B**) Spearman correlation between Log_2_FC in T cell abundance and age. (**C**) Absolute abundance of CD8^+^ and (**D**) CD4^+^ T cell subsets in patients stratified by age. Log_2_FC values are marked within joining segments. (**E**) CD8^+^ and (**F**) CD4^+^ T subset abundance for CMV^-^ and CMV^+^ patients as per (C) and (D). (**G**) Age-related differences in pre- to on-treatment log_2_FC in marker expression frequency by total CD8^+^ and CD4^+^ T cells. Balloon size represents -log_10_(*p*) and bold borders mark *p*<0.05. Colour represents delta log_2_FC (old - young); positive (red) values denote features with greater upregulation in older patients. (**H**) Log_2_FC in ICOS expression by CD8^+^ and CD4^+^ TemRO cells and (**I**) granzyme B by total CD4^+^ T and CD8^+^ TemRO cells between younger and older patients. (**J**) Log_2_FC in TNFα and TBET expression by total CD4^+^ T cells in CMV^-^ and CMV^+^patients. All plots show medians. *P* values from Mann Whitney tests unless otherwise noted.

## Discussion

We here determined the effect of CMV infection and age on the clinical and immunological response to immune checkpoint blockade in a cohort of patients with advanced melanoma and, consistent with most previous findings, we found no effect of age on response rate, overall survival, progression-free survival, or the occurrence of severe toxicity. Similarly, and despite recent observations that CMV seropositivity may improve overall survival following PD-1 blockade monotherapy in advanced melanoma patients (24), we found no effect of CMV status on clinical outcomes in our cohort.

Interestingly, we noted a cross-over survival curve pattern, precluding statistical analysis but suggesting that CMV^+^ patients had an early OS benefit before being disadvantaged after approximately 2-3 years. Supporting this, CMV^+^ patients receiving PD-1 blockade for advanced non-small cell lung cancer (NSCLC) did not have an OS benefit and had marginally worse PFS compared to CMV^-^ patients (25), suggesting that the association between CMV and ICB outcome may be cancer type-specific or inconsistent. We were unable to test associations between BRAF mutation status, CMV serostatus, and clinical outcomes within treatment groups due to limited patient numbers. Hence, larger pan-cancer studies will be required to determine if a robust overarching association exists between CMV and ICB treatment outcome. In accordance with recent work, we validated the presence of an immune-aged profile in patients ≥65 years of age, and a CMV-infected profile in seropositive patients (24,31). Importantly, we based our analyses on absolute immune cell counts rather than proportional frequencies, allowing us to accurately define the changes induced by these features. We demonstrated that CMV infection does not affect naive T cells as previously reported in melanoma patients (24), but solely drives expansion of functional, non-exhausted CD8^+^ TemRA and CD4^+^ TemRO and TemRA cells. In contrast, ageing was associated with a significant loss of naive CD8^+^ T cells and an increase in CD8^+^ TemRA cells. CD4^+^ T cells were differently affected with no change to naive cells but decreased Tcm, T_FH_, and T_H17_ subsets. These findings are largely consistent with a study assessing the impact of age and CMV on T cell subsets in healthy individuals using absolute numbers (65), and suggest that the effects of ageing and CMV are mostly independent and not altered by the presence of cancer. Ostensibly, an age-related decrease in baseline naive T cell abundance may impair responses to ICB, while CMV-induced effector memory expansion should support response (23,66,67). The lack of clinical impact observed would suggest that these peripheral immune features are not crucial for ICB efficacy or that the alterations observed were not large enough to influence clinical outcome.

We also noted significant declines in T_FH_ and B cells with age, as well as decreased ICOS expression on T_FH_. Decreases in total B cells were also reflected by a loss of Bmem, particularly IgD^+^IgM^+^ and IgG1^+^ subsets. Taken together, this may suggest that aged patients have diminished humoral immunity driven by an altered T-cell phenotype. This paradigm has been observed previously, with reduced vaccine responsiveness in older patients (13). Impaired vaccine efficacy was previously found to correlate with an increased frequency of CD21^low^ CD11c^+^ age-associated (atypical) B cells (atBC) in patients receiving ICB (68). Previous studies have identified an accumulation of these atBC in situations of repeated or chronic antigen stimulation in humans and in murine ageing models (69). atBC are also proportionally increased following ICB, where these have been associated with toxicity in melanoma patients or non-response in NSCLC patients (70,71). In contrast, we observed that neither age nor ICB altered the absolute abundance of atBCs. Rather, atBCs did not decline with age- or treatment as was observed for other B cell subsets, and so atBC were proportionally enriched in older patients and after ICB treatment. We here defined atBCs as CD21^low^ CD11c^+^ Bmem but note that the identification of these cells varies between studies, and their functional importance remains debated (72). Nonetheless, these results suggest that atBCs are stable throughout age and resistant to depletion during ICB treatment, and that impaired humoral immunity may be due to depletion of the conventional Bmem compartment or impaired T-cell help rather than accumulation of atBCs.

Whilst age and CMV had distinct effects on pre-treatment immunity, we observed that one cycle of ipi+nivo induced dramatic immune alterations, of which some were shared at a lesser magnitude following nivo/pembro or rela+nivo, consistent with previous findings (60,66,73–75). Age and CMV status had minimal impact on these changes, and so age- and CMV-associated phenotypes persisted during treatment. We suggest that the lack of impact of age and CMV infection on clinical outcomes may arise directly from their lack of impact on the immunological effects of ICB. The decline of naive cells with age that we observed did not significantly diminish the overall size of the T-cell compartment. Rather, memory T cells expanded to make up a greater proportion (76). An increased abundance of memory T cells has been associated with improved response to treatment in numerous studies (23,63,66,67,73), suggesting that the impact of a declining naive compartment on ICB efficacy may be balanced by the expansion of effector memory cells.

This study utilized a prospectively collected real-world cohort from a single institution and hence contains several limitations. Notably, a relatively small number of patients receiving nivo/pembro or rela+nivo were available for analysis, limiting our ability to evaluate the impact of age and CMV on the immune changes induced by these treatments, and no tumor data were available. Cohort size also prevented robust analysis of the interaction of age and CMV on immune phenotype. However, this work was strengthened overall by paired pre- and on-treatment data in patients with a uniform cancer type stratified by treatment, avoiding confounding due to cohort heterogeneity. Our analysis was also strengthened through the inclusion of absolute immune cell numbers per microliter of blood, preventing over-interpretation of proportional changes.

We here showed that while CMV seropositivity and ageing each had pronounced impacts on pre-treatment immune composition, neither meaningfully altered clinical or immunological response to treatment. Accordingly, the data presented here does not support the inclusion of these factors into clinical decision making.

## Data Availability

The data generated in this study are available upon request from the corresponding authors.

## Acknowledgements

We gratefully acknowledge the participants who donated samples for this study and the support of the Alfred Cancer Biobank. We also thank Alfred Research Alliance Flowcore staff for cytometry support, members of the Allergy and Clinical Immunology Laboratory for assistance with sample processing, Dr Gemma Hartley for production of recombinant HA protein, A/Prof Vivek Naranbhai for the comment that sparked this analysis, and Prof Mark Hogarth, Dr Bruce Wines, Sandra Esparon, and Reema Bajaj of the Burnet Institute for assistance with protein production.

## Authors’ contributions

Conceptualization: JME, SS, MCA, MCvZ

Data curation: JME, MCA

Formal analysis: JME

Investigation: JME

Methodology: JME, SS, MCA, MCvZ

Project administration: JME, SS, RS, HB, CO, MCA, MCvZ

Software: JME

Visualization: JME, MCA, MCvZ

Resources: SS, RS, HB, CO, MS, MCvZ

Supervision: SS, MCA, MCvZ

Writing – original draft: JME, MCA, MCvZ

Writing – review and editing: JME, SS, MCA, MCvZ

Funding acquisition: JME, SS, MS

