## Supplementary Tables and Figures for "Age and CMV are not associated with clinical or immunological response to immune checkpoint blockade"

### **Supplementary Tables** (n=8) **and Figures** (n=9)

**Supplementary Table 1**: TruCount antibody panel.

| **Target** | **Fluorochrome** | **Clone** | **Vendor** | **RRID^** | **Test use** | |
|---|---|---|---|---|---|---|
| | | | | | **µL/test** | **µg mL$^{-1}$** |
| CD3 | FITC | UCHT1 | BD Biosciences | AB_395739 | 3 | 0.56 |
| CD4 | BV605 | RPA-T4 | BD Biosciences | AB_2744420 | 0.5 | 0.36 |
| CD8 | BV510 | SKI | BD Biosciences | AB_2722546 | 0.5 | 1.43 |
| CD14 | APC-H7 | MφP9 | BD Biosciences | AB_1645725 | 1 | 0.36 |
| CD16 | PE | 3GA | Biolegend | AB_314208 | 0.7 | 1.00 |
| CD19 | PE-Cy7 | SJ25C1 | BD Biosciences | AB_396893 | 1 | 0.71 |
| CD45 | PerCP-Cy5.5 | 2D1 | BD Biosciences | AB_400194 | 4 | 0.34 |
| CD56 | PE | B159 | BD Biosciences | AB_395906 | 5 | 1.79 |
| HLA-DR | APC | L243 | Biolegend | AB_314688 | 2 | 0.71 |

^RRID: Research resource identifier

**Supplementary Table 2**: Resting and activated T cell flow cytometry reagents.

| Target | Fluorochrome | Clone | Supplier | RRID^ | Panel usage# | Test use | |
|---|---|---|---|---|---|---|---|
| | | | | | | µL/test | µg mL$^{-1}$ |
| CD3 | BUV805 | UCHT1 | BD Biosciences | AB_2800945 | 1,2 | 1.25 | 2.5 |
| CD4 | cFluor YG584 | SK3 | Cytek Biosciences | AB_2885083 | 1,2 | 1.25 | 0.5 |
| CD8 | Spark Blue 550 | SK1 | Biolegend | AB_2819982 | 1,2 | 0.25 | 0.1 |
| CD16 | BV510 | 3GA | BD Biosciences | AB_2819982 | 1 | 1.25 | 5 |
| CD19 | BUV563 | SJ25C1 | BD Biosciences | AB_2744296 | 1,2 | 0.3 | 0.6 |
| CD25 | PE-Fire700 | M-A251 | Biolegend | AB_2870202 | 1,2 | 2.5 | 2.5 |
| CD39 | APC-Fire750 | A1 | Biolegend | AB_2876678 | 1 | 0.5 | 0.5 |
| CD45 | PerCP | HI30 | Biolegend | AB_2650838 | 1,2 | 0.5 | 2 |
| CD45RA | BUV496 | HI100 | BD Biosciences | AB_893341 | 1,2 | 0.04 | 0.0064 |
| CD56 | BUV737 | B159 | BD Biosciences | AB_2874456 | 1 | 0.4 | 1.6 |
| CD57 | PB | HNK-1 | Biolegend | AB_2871176 | 1 | 0.2 | 0.2 |
| CD95 | PE-Dazzle594 | DX2 | Biolegend | AB_2562458 | 1,2 | 0.625 | 1.25 |
| CD127 | R718 | HIL-7R-M21 | BD Biosciences | AB_2564221 | 1,2 | 2.5 | 2.5 |
| CCR7 | BV750 | GO43H7 | Biolegend | AB_2869977 | 1,2 | 2.5 | 10 |
| CTLA-4 | BV785 | BNI3 | Biolegend | AB_2810582 | 2 | 0.6 | 1.2 |
| CXCR5 | BV605 | RF8B2 | BD Biosciences | AB_2740110 | 1 | 2.5 | 10 |
| EOMES | PE-Cy7 | WD1928 | ThermoFisher | AB_2573456 | 1 | 2.5 | 0.6 |
| GrzmB | BV510 | GB11 | BD Biosciences | AB_2738174 | 2 | 0.3 | 1.2 |
| ICOS | BUV661 | DX29 | BD Biosciences | AB_2871056 | 1 | 1.25 | 5 |
| IFNγ | BV605 | B27 | BD Biosciences | AB_2737926 | 2 | 0.6 | 1.2 |
| IL-2 | AF488 | MQ1-17H12 | BD Biosciences | AB_2738566 | 2 | 2.5 | 2.5 |
| IL-4 | APC | MP4-25D2 | Biolegend | AB_2295923 | 2 | 2.5 | 0.2 |
| IL-10 | BV421 | JES3-9D7 | BD Biosciences | AB_493368 | 2 | 2.5 | 2.5 |
| IL-17A | PE-Cy7 | BL168 | Biolegend | AB_315131 | 2 | 0.2 | 0.6 |
| IRF4 | AF647 | IRF4.3E4 | Biolegend | AB_2564047 | 1 | 1.25 | 2.5 |
| Ki67 | BUV395 | B59 | BD Biosciences | AB_2738577 | 1 | 1.25 | 7.5 |
| KLRG1 | APC | 2F1 | Biolegend | AB_10645509 | 1 | 0.3 | 0.3 |
| PD-1 | BV786 | EH12.1 | BD Biosciences | AB_2738425 | 1 | 1.25 | 5 |
| Tbet | PE-Cy5 | 4B10 | ThermoFisher | AB_2815071 | 1 | 0.4 | 1.6 |
| TCRγδ | PerCP-Vio700 | REA591 | Miltenyi Biotec | AB_2733074 | 1,2 | 1 | 8 |
| TIGIT | BV421 | A5153G | Biolegend | AB_2632925 | 1 | 2.5 | 2.5 |
| TIM-3 | BB515 | 7D3 | BD Biosciences | AB_2744368 | 1 | 2.5 | 5 |
| TNFα | BUV395 | MAb11 | BD Biosciences | AB_2738533 | 2 | 0.5 | 2 |
| TOX | PE | TXRX10 | ThermoFisher | AB_10855034 | 1 | 0.625 | 2.5 |
| Viability | Live/Dead Blue | L34962 | ThermoFisher | *NA* | 1,2 | 1.25 | *NA* |

^Research resource identifier. #Panel 1: resting; panel 2: activated.

**Supplementary Table 3**: B cell antibody panel.

| Target | Fluorochrome | Clone | Supplier | RRID^ | Strep control | Test use | |
|---|---|---|---|---|---|---|---|
| | | | | | | µL/test | µg mL$^{-1}$ |
| CD3 | BUV805 | UCHT1 | BD | AB_2870183 | Yes | 6.25 | 2.5 |
| CD11c | PE-Vio 670 | REA618 | Miltenyi | AB_3663134 | No | 2.5 | 0.25 |
| CD19 | BUV563 | SJ25C1 | BD | AB_2870202 | Yes | 1.5 | 0.6 |
| CD21 | BV711 | B-ly4 | BD | AB_2738040 | No | 6.25 | 1.25 |
| CD27 | RB744 | M-T271 | BD | AB_3685904 | Yes | 6.25 | 5 |
| CD38 | APC-R700 | HIT2 | BD | AB_2744373 | No | 6.25 | 2.5 |
| CD71 | BV786 | M-A712 | BD | AB_2738414 | No | 2.5 | 0.12 |
| IgA | PE-Vio 770 | REA1014 | Miltenyi | AB_2727741 | No | 1.25 | 0.375 |
| IgD | APC-Fire 750 | W18340F | Biolegend | AB_3097320 | Yes | 2.5 | 2 |
| IgG1 | PE | G17-1 | BD custom | NA | No | 0.25 | 0.2 |
| IgG2 | FITC | SAG2 | BD | AB_3720082 | No | 5 | 0.66 |
| IgG2 | PE | SAG2 | BD | AB_3720081 | No | 5 | 1 |
| IgG3 | FITC | HP6047 | BD custom | NA | No | 1.25 | 1 |
| IgG4 | APC | SAG4 | BD | AB_3720080 | No | 5 | 0.84 |
| IgM | Pacific Blue | MHM-88 | Biolegend | AB_10574307 | No | 0.5 | 1 |
| PD-L1 | RB780 | MIH1 | BD Opti | AB_3687837 | No | 12.5 | 10 |
| TIM-1 | BUV737 | 1D12 | BD Opti | AB_2872895 | No | 12.5 | 10 |
| Viability | Live/Dead Blue | L34962 | ThermoFisher | NA | Yes | 6.25 | NA |
| Streptavidin | BUV615 | NA | BD | NA | Yes | Variable | Variable |
| Streptavidin | BV650 | NA | Biolegend | NA | Yes | Variable | Variable |

^Research resource identifier

**Supplementary Table 4:** Resting T cell panel population definitions.

| | Population name | Phenotype definition |
|---|---|---|
| | *All populations gated from live lymphocytes:* CD45$^{+}$ SSC$^{low}$ Live/Dead Blue$^{-}$ | |
| 1 | B cells | CD3$^{-}$ CD19$^{+}$ |
| 2 | NK cells | CD3$^{-}$ CD19$^{-}$ CD16/CD56$^{+}$ |
| 3 | T cells | CD3$^{+}$ CD19$^{-}$ |
| 4 | ∟TCRγδ$^{+}$ | CD3$^{+}$ CD19$^{-}$ TCRγδ$^{+}$ |
| 7 | ∟TCRαβ$^{+}$ | CD3$^{+}$ CD19$^{-}$ TCRγδ$^{-}$ |
| 10 | ∟CD4$^{-}$ CD8$^{-}$ | CD3$^{+}$ CD19$^{-}$ TCRγδ$^{-}$ CD4$^{-}$ CD8$^{-}$ |
| 11 | ∟CD4 T cells | CD3$^{+}$ CD19$^{-}$ TCRγδ$^{-}$ CD4$^{+}$ CD8$^{-}$ |
| 12 | ∟Treg | CD3$^{+}$ CD19$^{-}$ TCRγδ$^{-}$ CD4$^{+}$ CD25$^{+}$ CD127$^{-/lo}$ |
| 13 | ∟NOT Treg | CD3$^{+}$ CD19$^{-}$ TCRγδ$^{-}$ CD4$^{+}$ CD25$^{+/lo}$ CD127$^{+/-}$ |
| 14 | ∟Naive/stem-like | CD3$^{+}$ CD19$^{-}$ TCRγδ$^{-}$ CD4$^{+}$ CD25$^{+/lo}$ CD127$^{+/-}$ CD45RA$^{+}$ CCR7$^{+}$ |
| 15 | ∟Tnaive *(TN)* | CD3$^{+}$ CD19$^{-}$ TCRγδ$^{-}$ CD4$^{+}$ CD25$^{+/lo}$ CD127$^{+/-}$ CD45RA$^{+}$ CCR7$^{+}$ CD95$^{-}$ |
| 16 | ∟Tscm *(TS$_{B}$M$_{p}$)* | CD3$^{+}$ CD19$^{-}$ TCRγδ$^{-}$ CD4$^{+}$ CD25$^{+/lo}$ CD127$^{+/-}$ CD45RA$^{+}$ CCR7$^{+}$ CD95$^{+}$ |
| 17 | ∟Tcm *(TS$_{B}$M)* | CD3$^{+}$ CD19$^{-}$ TCRγδ$^{-}$ CD4$^{+}$ CD25$^{+/lo}$ CD127$^{+/-}$ CD45RA$^{-}$ CCR7$^{+}$ |
| 18 | ∟TemRO *(TD$_{B}$M)* | CD3$^{+}$ CD19$^{-}$ TCRγδ$^{-}$ CD4$^{+}$ CD25$^{+/lo}$ CD127$^{+/-}$ CD45RA$^{-}$ CCR7$^{-}$ |
| 19 | ∟TemRA *(CD45RA$^{+}$ TD$_{B}$M)* | CD3$^{+}$ CD19$^{-}$ TCRγδ$^{-}$ CD4$^{+}$ CD25$^{+/lo}$ CD127$^{+/-}$ CD45RA$^{+}$ CCR7$^{-}$ |
| 20 | ∟Tfh | CD3$^{+}$ CD19$^{-}$ TCRγδ$^{-}$ CD4$^{+}$ CD25$^{+/lo}$ CD127$^{+/-}$ CD45RA$^{+}$ CXCR5$^{+}$ |
| 21 | ∟CD8 T cells | CD3$^{+}$ CD19$^{-}$ TCRγδ$^{-}$ CD8$^{+}$ CD4$^{-}$ |
| 22 | ∟Naive/stem-like | CD3$^{+}$ CD19$^{-}$ TCRγδ$^{-}$ CD8$^{+}$ CD45RA$^{+}$ CCR7$^{+}$ |
| 23 | ∟Tnaive *(TN)* | CD3$^{+}$ CD19$^{-}$ TCRγδ$^{-}$ CD8$^{+}$ CD25$^{+/lo}$ CD127$^{+/-}$ CD45RA$^{+}$ CCR7$^{+}$ CD95$^{-}$ |
| 24 | ∟Tscm *(TS$_{B}$M$_{p}$)* | CD3$^{+}$ CD19$^{-}$ TCRγδ$^{-}$ CD8$^{+}$ CD25$^{+/lo}$ CD127$^{+/-}$ CD45RA$^{+}$ CCR7$^{+}$ CD95$^{+}$ |
| 25 | ∟Tcm *(TS$_{B}$M)* | CD3$^{+}$ CD19$^{-}$ TCRγδ$^{-}$ CD8$^{+}$ CD45RA$^{-}$ CCR7$^{+}$ |
| 26 | ∟TemRO *(TD$_{B}$M)* | CD3$^{+}$ CD19$^{-}$ TCRγδ$^{-}$ CD8$^{+}$ CD45RA$^{-}$ CCR7$^{-}$ |
| 27 | ∟TemRA *(CD45RA$^{+}$ TD$_{B}$M)* | CD3$^{+}$ CD19$^{-}$ TCRγδ$^{-}$ CD8$^{+}$ CD45RA$^{+}$ CCR7$^{-}$ |

**Supplementary Table 5**: Activated T cell panel population definitions.

| | Population name | Phenotype definition |
|---|---|---|
| | *All populations gated from live lymphocytes:* $CD45^{+}$ $SSC^{low}$ Live/Dead $Blue^{-}$ | |
| 1 | T + NK cells | $CD19^{-}$ |
| 2 | ∟ $TCR\gamma\delta^{+}$ | $CD19^{-}$ $TCR\gamma\delta^{+}$ |
| 3 | ∟ $TCR\gamma\delta^{-}$ | $CD19^{-}$ $TCR\gamma\delta^{-}$ |
| 4 | ∟ CD4 T cells | $CD19^{-}$ $TCR\gamma\delta^{-}$ $CD4^{+}$ $CD8^{-}$ |
| 5 | ∟ Treg | $CD19^{-}$ $TCR\gamma\delta^{-}$ $CD4^{+}$ $CD25^{+}$ $CD127^{-/lo}$ |
| 6 | ∟ NOT Treg | $CD19^{-}$ $TCR\gamma\delta^{-}$ $CD4^{+}$ $CD25^{+/lo}$ $CD127^{+/-}$ |
| 7 | ∟ Naive/stem-like | $CD19^{-}$ $TCR\gamma\delta^{-}$ $CD4^{+}$ $CD25^{+/lo}$ $CD127^{+/-}$ $CD45RA^{+}$ $CCR7^{+}$ |
| 8 | ∟ Tnaive *(TN)* | $CD19^{-}$ $TCR\gamma\delta^{-}$ $CD4^{+}$ $CD25^{+/lo}$ $CD127^{+/-}$ $CD45RA^{+}$ $CCR7^{+}$ $CD95^{-}$ |
| 9 | ∟ Tscm *($TS_BM_p$)* | $CD19^{-}$ $TCR\gamma\delta^{-}$ $CD4^{+}$ $CD25^{+/lo}$ $CD127^{+/-}$ $CD45RA^{+}$ $CCR7^{+}$ $CD95^{+}$ |
| 10 | ∟ Tcm *($TS_BM$)* | $CD19^{-}$ $TCR\gamma\delta^{-}$ $CD4^{+}$ $CD25^{+/lo}$ $CD127^{+/-}$ $CD45RA^{-}$ $CCR7^{+}$ |
| 11 | ∟ TemRO *($TD_BM$)* | $CD19^{-}$ $TCR\gamma\delta^{-}$ $CD4^{+}$ $CD25^{+/lo}$ $CD127^{+/-}$ $CD45RA^{-}$ $CCR7^{-}$ |
| 12 | ∟ TemRA *($CD45RA^{+}$ $TD_BM$)* | $CD19^{-}$ $TCR\gamma\delta^{-}$ $CD4^{+}$ $CD25^{+/lo}$ $CD127^{+/-}$ $CD45RA^{+}$ $CCR7^{-}$ |
| 13 | ∟ Th1 | $CD19^{-}$ $TCR\gamma\delta^{-}$ $CD4^{+}$ $CD25^{+/lo}$ $CD127^{+/-}$ $IL\text{-}17A^{-}$ $IFN\gamma^{+}$ |
| 14 | ∟ Th2 | $CD19^{-}$ $TCR\gamma\delta^{-}$ $CD4^{+}$ $CD25^{+/lo}$ $CD127^{+/-}$ $IL\text{-}17A^{-}$ $IFN\gamma^{-}$ $IL\text{-}4^{+}$ |
| 15 | ∟ Th17 | $CD19^{-}$ $TCR\gamma\delta^{-}$ $CD4^{+}$ $CD25^{+/lo}$ $CD127^{+/-}$ $IL\text{-}17A^{+}$ $IFN\gamma^{-}$ |
| 16 | ∟ CD8 T cells | $CD19^{-}$ $TCR\gamma\delta^{-}$ $CD8^{+}$ $CD4^{-}$ |
| 17 | ∟ Naive/stem-like | $CD19^{-}$ $TCR\gamma\delta^{-}$ $CD8^{+}$ $CD25^{+/lo}$ $CD127^{+/-}$ $CD45RA^{+}$ $CCR7^{+}$ |
| 18 | ∟ Tnaive *(TN)* | $CD19^{-}$ $TCR\gamma\delta^{-}$ $CD8^{+}$ $CD25^{+/lo}$ $CD127^{+/-}$ $CD45RA^{+}$ $CCR7^{+}$ $CD95^{-}$ |
| 19 | ∟ Tscm *($TS_BM_p$)* | $CD19^{-}$ $TCR\gamma\delta^{-}$ $CD8^{+}$ $CD25^{+/lo}$ $CD127^{+/-}$ $CD45RA^{+}$ $CCR7^{+}$ $CD95^{+}$ |
| 20 | ∟ Tcm *($TS_BM$)* | $CD19^{-}$ $TCR\gamma\delta^{-}$ $CD8^{+}$ $CD45RA^{-}$ $CCR7^{-}$ |
| 21 | ∟ TemRO *($TD_BM$)* | $CD19^{-}$ $TCR\gamma\delta^{-}$ $CD8^{+}$ $CD45RA^{-}$ $CCR7^{-}$ |
| 22 | ∟ TemRA *($CD45RA^{+}$ $TD_BM$)* | $CD19^{-}$ $TCR\gamma\delta^{-}$ $CD8^{+}$ $CD45RA^{+}$ $CCR7^{-}$ |

**Supplementary Table 6:** TruCount panel population definitions.

| | Population name | Phenotype definition |
|---|---|---|
| 1 | Granulocytes | $SSC^{high}$ $CD45^{+}$ |
| 2 | ∟neutrophils | $SSC^{high}$ $CD45^{+}$ $CD16/56^{+}$ |
| 3 | ∟eosinophils | $SSC^{high}$ $CD45^{+}$ $CD16/56^{-}$ |
| | Mononuclear cells | $SSC^{inter}$ $CD45^{+}$ |
| 5 | ∟M-DC | $SSC^{inter}$ $CD45^{+}$ $CD3^{-}$ $CD19^{-}$ $HLA\text{-}DR^{+}$ $CD16/56^{-}$ $CD14^{-}$ |
| 6 | ∟Monocytes | $SSC^{inter}$ $CD45^{+}$ $CD3^{-}$ $CD19^{-}$ $HLA\text{-}DR^{+}$ $CD16/56^{+/-}$ $CD14^{+/-}$ |
| 7 | ∟classical | $SSC^{inter}$ $CD45^{+}$ $CD3^{-}$ $CD19^{-}$ $HLA\text{-}DR^{+}$ $CD16/56^{-}$ $CD14^{+}$ |
| 8 | ∟intermediate | $SSC^{inter}$ $CD45^{+}$ $CD3^{-}$ $CD19^{-}$ $HLA\text{-}DR^{+}$ $CD16/56^{+}$ $CD14^{+}$ |
| 9 | ∟non-classical | $SSC^{inter}$ $CD45^{+}$ $CD3^{-}$ $CD19^{-}$ $HLA\text{-}DR^{+}$ $CD16/56^{+}$ $CD14^{-}$ |
| 10 | Lymphocytes | $SSC^{low}$ $CD45^{+}$ |
| 11 | ∟B cells | $SSC^{low}$ $CD45^{+}$ $CD3^{-}$ $CD19^{+}$ |
| 12 | ∟NK cells | $SSC^{low}$ $CD45^{+}$ $CD3^{-}$ $CD16/56^{+}$ |
| 13 | ∟T cells | $SSC^{low}$ $CD45^{+}$ $CD19^{-}$ $CD3^{+}$ |
| 14 | ∟$CD4^{+}$ T cells | $SSC^{low}$ $CD45^{+}$ $CD19^{-}$ $CD3^{+}$ $CD4^{+}$ $CD8^{-}$ |
| 15 | ∟$CD8^{+}$ T cells | $SSC^{low}$ $CD45^{+}$ $CD19^{-}$ $CD3^{+}$ $CD4^{-}$ $CD8^{+}$ |

**Supplementary Table 7**: B cell population definitions.

| | Population name | Phenotype definition |
|---|---|---|
| | *All populations gated from live lymphocytes:* | $FSC^{mid}SSC^{low}$Live/Dead $Blue^{-}$ |
| 1 | B cells | $CD3^{-}$ $CD19^{+}$ |
| 2 | ∟ Transitional | $CD3^{-}$ $CD19^{+}$ $CD38^{high}$ $CD27^{-}$ |
| 3 | ∟ Plasmablast | $CD3^{-}$ $CD19^{+}$ $CD38^{high}$ $CD27^{+}$ |
| 4 | ∟ Mature B | $CD3^{-}$ $CD19^{+}$ $CD38^{low/mid}$ $CD27^{+/-}$ |
| 5 | ∟ Naive mature | $CD3^{-}$ $CD19^{+}$ $CD38^{low/mid}$ $CD27^{-}$ $IgD^{+}$ |
| 6 | ∟ Memory (Bmem) | $CD3^{-}$ $CD19^{+}$ $CD38^{low/mid}$ $CD27^{+/-}$ $IgD^{+/-}$ |
| 7 | ∟ $IgD^{+}$ | $CD3^{-}$ $CD19^{+}$ $CD38^{low/mid}$ $CD27^{+/-}$ $IgD^{+}$ $IgM^{-}$ |
| 8 | ∟ $IgD^{+}$ $IgM^{+}$ | $CD3^{-}$ $CD19^{+}$ $CD38^{low/mid}$ $CD27^{+/-}$ $IgD^{+}$ $IgM^{+}$ |
| 9 | ∟ $IgM^{+}$ | $CD3^{-}$ $CD19^{+}$ $CD38^{low/mid}$ $CD27^{+/-}$ $IgD^{-}$ $IgM^{+}$ |
| 10 | ∟ Class switched | $CD3^{-}$ $CD19^{+}$ $CD38^{low/mid}$ $CD27^{+/-}$ $IgD^{-}$ $IgM^{-}$ |
| 11 | ∟ $IgG1^{+}$ | $CD3^{-}$ $CD19^{+}$ $CD38^{low/mid}$ $CD27^{+/-}$ $IgD^{-}$ $IgM^{-}$ $IgG1^{+}$ $IgG2^{-}$ $IgG3^{-}$ |
| 12 | ∟ $IgG2^{+}$ | $CD3^{-}$ $CD19^{+}$ $CD38^{low/mid}$ $CD27^{+/-}$ $IgD^{-}$ $IgM^{-}$ $IgG1^{-}$ $IgG2^{+}$ $IgG3^{-}$ |
| 13 | ∟ $IgG3^{+}$ | $CD3^{-}$ $CD19^{+}$ $CD38^{low/mid}$ $CD27^{+/-}$ $IgD^{-}$ $IgM^{-}$ $IgG1^{-}$ $IgG2^{-}$ $IgG3^{+}$ |
| 14 | ∟ IgG4 | $CD3^{-}$ $CD19^{+}$ $CD38^{low/mid}$ $CD27^{+/-}$ $IgD^{-}$ $IgM^{-}$ $IgG1^{-}$ $IgG2^{-}$ $IgG3^{-}$ $IgG4^{+}$ $IgA^{-}$ |
| 15 | ∟ IgA | $CD3^{-}$ $CD19^{+}$ $CD38^{low/mid}$ $CD27^{+/-}$ $IgD^{-}$ $IgM^{-}$ $IgG1^{-}$ $IgG2^{-}$ $IgG3^{-}$ $IgG4^{-}$ $IgA^{+}$ |
| 16 | ∟ Unclassified | $CD3^{-}$ $CD19^{+}$ $CD38^{low/mid}$ $CD27^{+/-}$ $IgD^{-}$ $IgM^{-}$ $IgG1^{-}$ $IgG2^{-}$ $IgG3^{-}$ $IgG4^{-}$ $IgA^{-}$ |
| 17 | ∟ Atypical | $CD3^{-}$ $CD19^{+}$ $CD38^{low/mid}$ $CD27^{+/-}$ $IgD^{+/-}$ $CD21^{-}$ $CD11c^{+}$ |

**Supplementary Table 8**: Cohort characteristics grouped by CMV serostatus.

| | **CMV- (n=28)** | **CMV+ (n=51)** | ***p*-value[#]** |
|---|---|---|---|
| Age (years) | | | |
| Median (range) | 66 (26-88) | 71 (25-92) | 0.33 |
| Sex | | | |
| Male | 22 (79%) | 35 (69%) | 0.44 |
| Female | 6 (21%) | 16 (31%) | |
| Best overall response (RECIST v1.1)[‡] | | | |
| Responder (PR, CR) | 18 (64%) | 33 (65%) | >0.99 |
| Non-responder (SD, PD, death) | 10 (36%) | 18 (35%) | |
| Toxicity (CTCAE v5.0[‡]) | | | |
| Any grade | 23 (82%) | 42 (82%) | >0.99 |
| Grade 3+ | 12 (43%) | 20 (39%) | 0.81 |
| Median time (days) to onset of severe toxicity (range) | 48 (14-98) | 46.5 (19-198) | 0.62 |
| Treatment | | | |
| Nivo/pembro | 8 (28%) | 14 (27%) | >0.99 |
| Ipi+nivo | 17 (61%) | 30 (59%) | |
| Rela+nivo | 3 (11%) | 7 (14%) | |
| Genomics | | | |
| *BRAF* V600 mutant | 6 (21%) | 13 (25%) | 0.64 |
| *NRAS* mutant | 12 (43%) | 17 (33%) | |
| *BRAF*/*NRAS* wt | 10 (36%) | 19 (37%) | |
| Not measured | 0 (0%) | 2 (4%) | |
| Stage of disease (AJCC 8th Ed.[‡]) | | | |
| IIIB | 1 (4%) | 1 (2%) | 0.48 |
| IIIC | 1 (4%) | 7 (14%) | |
| IIID | 1 (4%) | 1 (2%) | |
| IVA | 2 (7%) | 7 (14%) | |
| IVB | 3 (11%) | 8 (16%) | |
| IVC | 9 (32%) | 16 (31%) | |
| IVD | 11 (39%) | 11 (22%) | |
| Metastatic disease sites | | | |
| Brain | 11 (39%) | 11 (22%) | 0.12 |
| Liver | 8 (29%) | 9 (18%) | 0.27 |
| Multi-site involvement (≥3 sites) | 16 (57%) | 17 (33%) | 0.057 |
| Baseline LDH | | | |
| Elevated (>upper limit of normal) | 11 (39%) | 17 (33%) | 0.87 |
| Normal | 12 (43%) | 23 (45%) | |
| Not measured | 5 (18%) | 11 (22%) | |
| ECOG score[‡] | | | |
| 0 | 18 (64%) | 26 (51%) | 0.66 |
| 1 | 10 (36%) | 22 (43%) | |
| 2 | 0 (0%) | 2 (4%) | |
| Unknown | 0 (0%) | 1 (2%) | |
| Prior therapy | | | |
| Treatment naive | 18 (64%) | 31 (61%) | 0.81 |
| Immunotherapy | 6 (21%) | 11 (22%) | >0.99 |
| Targeted therapy | 2 (7%) | 4 (8%) | >0.99 |
| Radiotherapy | 2 (7%) | 11 (22%) | 0.12 |
| Cytotoxic | 0 (0%) | 1 (2%) | NA |

**Supplementary Table 9**: Cohort characteristics grouped by age.

| | **Young (<65 years) n=30** | **Old (≥65 years) n=49** | ***p*-value[#]** |
|---|---|---|---|
| Sex | | | |
| Male | 20 (67%) | 37 (76%) | 0.44 |
| Female | 10 (33%) | 12 (24%) | |
| Best overall response (RECIST v1.1)[‡] | | | |
| Responder (PR, CR) | 18 (60%) | 33 (67%) | 0.63 |
| Non-responder (SD, PD, death) | 12 (40%) | 16 (33%) | |
| Toxicity (CTCAE v5.0[‡]) | | | |
| Any grade | 25 (83%) | 40 (82%) | >0.9999 |
| Grade 3+ | 15 (50%) | 17 (35%) | 0.24 |
| Median time (days) to onset of severe toxicity (range) | 43 (14-198) | 54 (19-127) | 0.054 |
| Treatment | | | |
| Nivo/pembro | 4 (13%) | 18 (37%) | **0.018** |
| Ipi+nivo | 24 (80%) | 23 (47%) | |
| Rela+nivo | 2 (7%) | 8 (16%) | |
| Genomics | | | |
| *BRAF* V600 mutant | 10 (33%) | 9 (18%) | 0.44 |
| *NRAS* mutant | 10 (33%) | 20 (41%) | |
| *BRAF*/*NRAS* wt | 10 (33%) | 18 (37%) | |
| Not measured | 0 (0%) | 2 (4%) | |
| Stage of disease (AJCC 8th Ed.[‡]) | | | |
| IIIB | 0 (0%) | 2 (4%) | 0.72 |
| IIIC | 3 (10%) | 5 (10%) | |
| IIID | 1 (3%) | 1 (2%) | |
| IVA | 4 (13%) | 5 (10%) | |
| IVB | 6 (20%) | 5 (10%) | |
| IVC | 7 (23%) | 18 (37%) | |
| IVD | 9 (30%) | 13 (27%) | |
| Metastatic disease sites | | | |
| Brain | 9 (30%) | 13 (27%) | 0.8 |
| Liver | 4 (13%) | 14 (29%) | 0.17 |
| Multi-site involvement (≥3 sites) | 11 (37%) | 22 (45%) | 0.49 |
| Baseline LDH | | | |
| Elevated (>upper limit of normal) | 13 (43%) | 15 (31%) | 0.37 |
| Normal | 13 (43%) | 22 (45%) | |
| Not measured | 4 (13%) | 12 (24%) | |
| ECOG score[‡] | | | |
| 0 | 21 (70%) | 23 (47%) | **0.039** |
| 1 | 8 (27%) | 24 (49%) | |
| 2 | 0 (0%) | 2 (4%) | |
| Unknown | 1 (3%) | 0 (0%) | |
| CMV serostatus | | | |
| Positive | 18 (60%) | 33 (67%) | 0.63 |
| Prior therapy | | | |
| Treatment naive | 18 (67%) | 33 (67%) | 0.63 |
| Immunotherapy | 11 (37%) | 8 (16%) | 0.058 |
| Targeted therapy | 4 (13%) | 2 (4%) | 0.19 |
| Radiotherapy | 5 (17%) | 6 (12%) | 0.74 |
| Cytotoxic | 0 (0%) | 1 (2%) | NA |

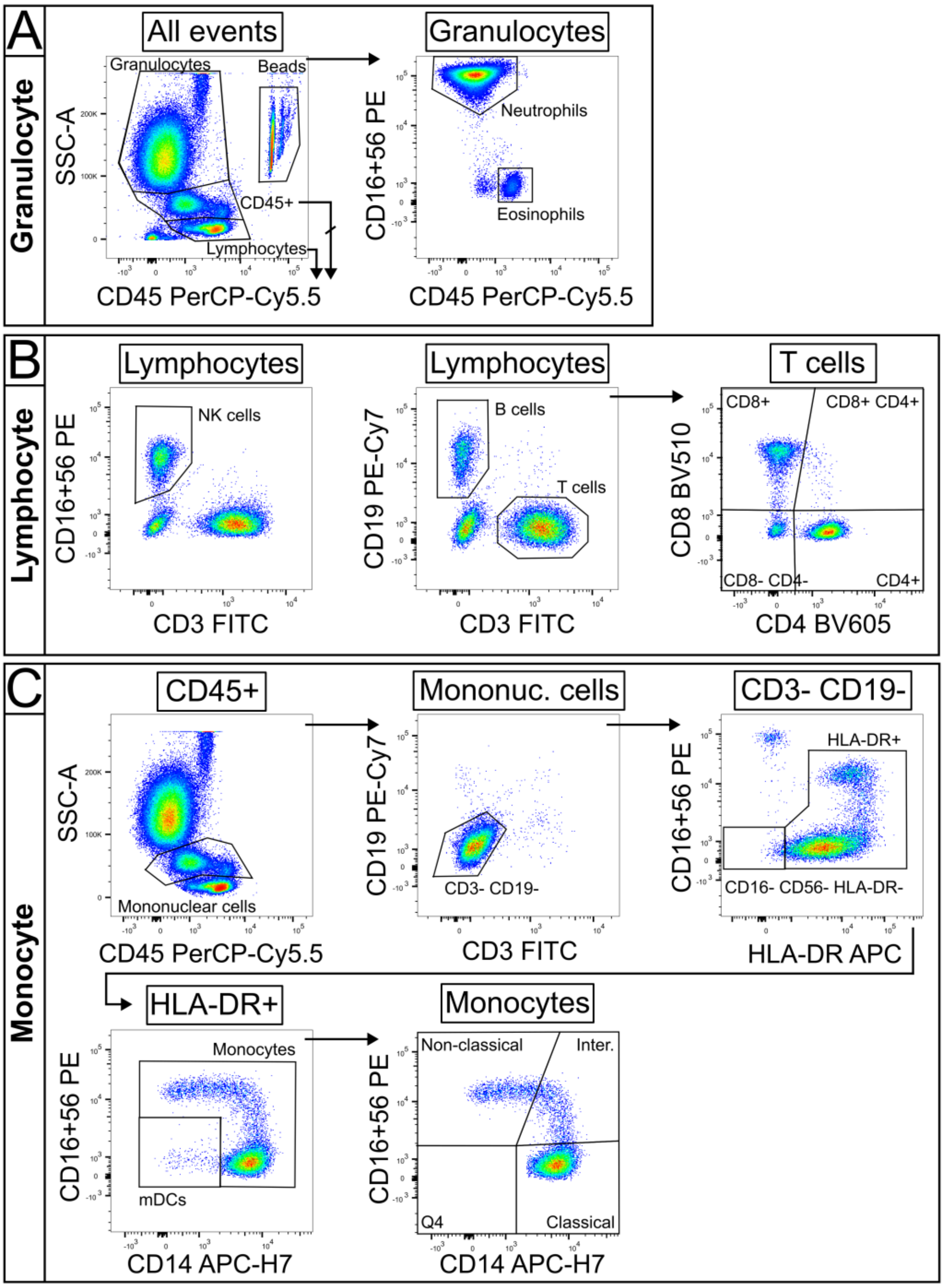


**Supplementary Figure 1**: Gating scheme for TruCount analysis of whole blood.

Whole blood was stained in a Lyse/No Wash protocol and analyzed on either an LSR II or FACSLyric (BD Biosciences). For FACSLyric acquisition (pictured), a CD45 threshold was set to exclude debris. (A) Major leukocyte populations were gated based on CD45 expression; granulocytes (CD45$^{+}$ SSC$^{hi}$), lymphocytes (CD45$^{+}$ SSC$^{lo}$) and TruCount beads (CD45$^{hi}$), and granulocytes were separated into neutrophils (CD16$^{+}$) and eosinophils (CD16$^{-}$). (B) Lymphocytes were separated into NK cells (CD3$^{-}$ CD16/56$^{+}$), B cells (CD3$^{-}$ CD19$^{+}$) and T cells (CD3$^{+}$ CD19$^{-}$). T cells were further characterized based on CD4 and CD8 expression. (C) Mononuclear cells were sequentially gated to remove contaminating cells and separated into myeloid dendritic cells (mDC; CD16/56$^{-}$ CD14$^{-}$ HLA-DR$^{+}$) and monocytes (HLA-DR$^{+}$). Monocytes were classified as classical (CD14$^{+}$ CD16/56$^{-}$), intermediate (CD14$^{+}$ CD16/56$^{+}$) and non-classical (CD14$^{-}$ CD16/56$^{+}$). HLA-DR expression heatmap overlay was used to assist monocyte subset gating (not shown).

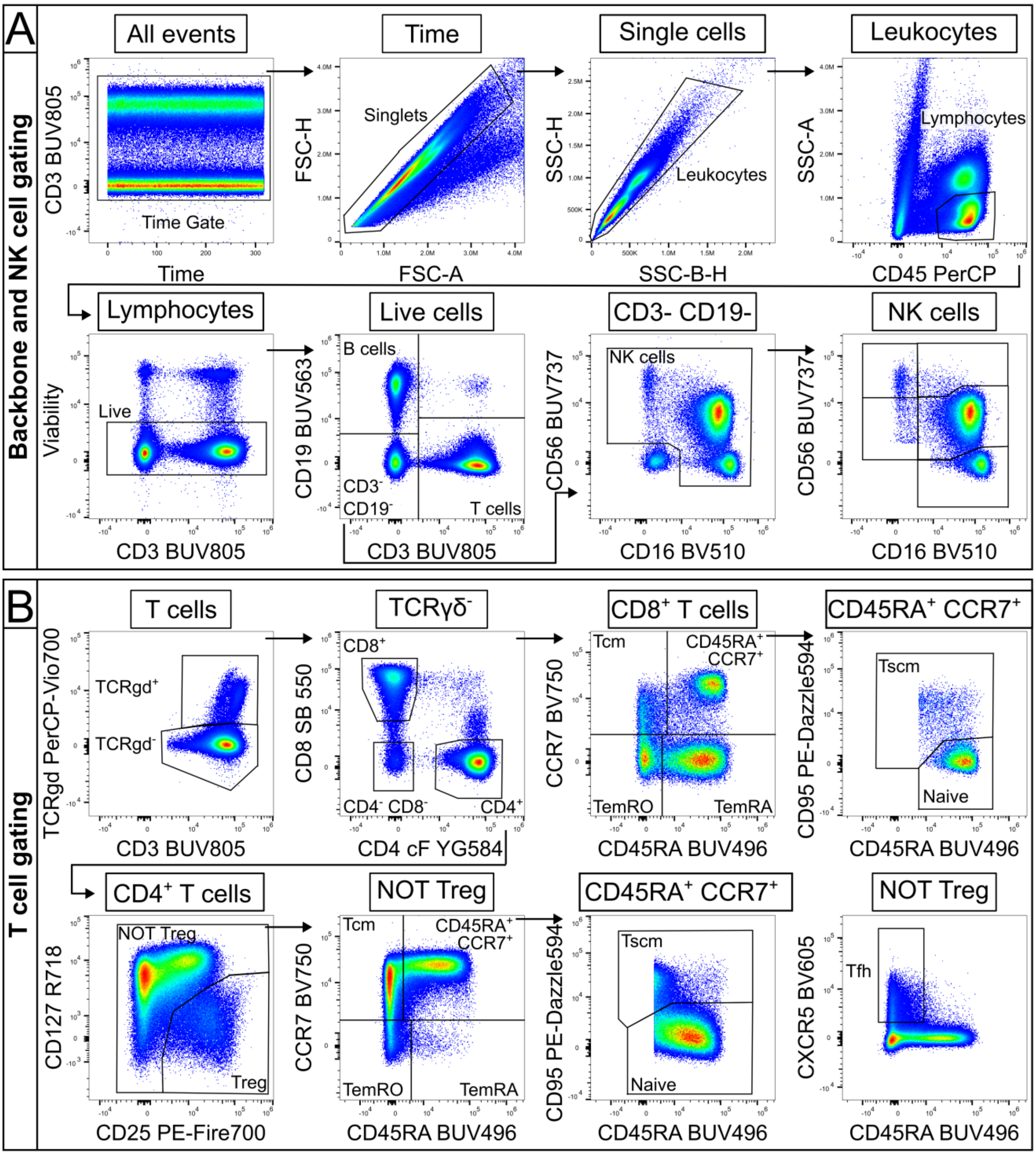


**Supplementary Figure 2**. Gating scheme for the 27-color resting T cell panel.

(A) Single, live lymphocytes were gated sequentially before division into B cells (CD19+), T cells (CD3+) and NK cells (CD19- CD3- CD16/56+). NK-cell subsets were further separated based on differential expression of CD16 and CD56. (B) TCRγδ+ T cells were defined before CD4 and CD8 lineage gating on the TCRγδ- subset. CD4+ Treg cells (CD25+ CD127-/lo) were excluded before gating of memory populations from the NOT Treg gate. Memory populations for both CD4+ and CD8+ T cells were defined as naive (CD45RA+ CCR7+ CD95lo), stem cell-like memory (Tscm; CD45RA+ CCR7+ CD95hi), central memory (Tcm; CD45RA- CCR7+), Effector Memory CD45RO (TemRO; CD45RA- CCR7-) and effector memory CD45RA (TemRA; CD45RA+ CCR7-). Circulating CD4+ Tfh cells were defined as CD45RA- CXCR5+.

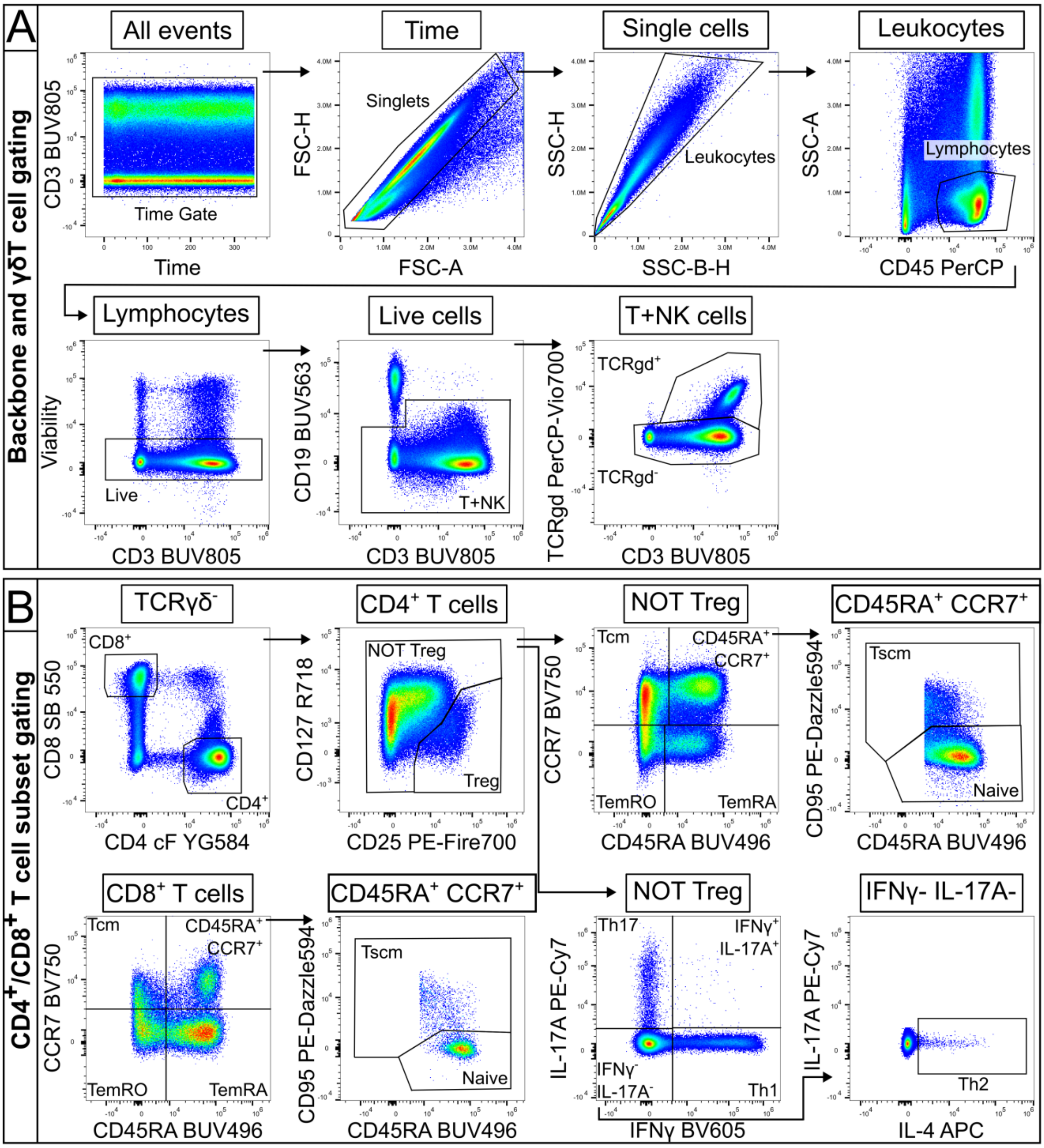


**Supplementary Figure 3.** Gating scheme for the 20-color CD3+CD28-stimulated T cell panel.

(A) Events were gated sequentially to select single, live lymphocytes. Due to CD3 down-regulation in some samples, T cells were gated from a combined T and NK cell gate ($CD19^{-}$ $CD3^{+/-}$) before gating of $TCR\gamma\delta^{+}$ cells. (B) $CD4^{+}$ and $CD8^{+}$ T cell subsets were gated as $CD19^{-}$ $TCR\gamma\delta^{-}$ $CD4^{+}$ or $CD19^{-}$ $TCR\gamma\delta^{-}$ $CD8^{hi}$, respectively (NK cells are $TCR\gamma\delta^{-}$ $CD8^{-/lo}$ $CD4^{-}$). Treg were gated as $CD25^{+}$ $CD127^{-/lo}$, with subsequent $CD4^{+}$ T memory and Th-cell gating from the 'NOT Treg' gate. Th cell subsets were defined by canonical cytokine expression (Th1, $IFN\gamma^{+}$; Th17, IL-$17A^{+}$; and Th2, $IFN\gamma^{-}$ IL-$17A^{-}$ IL-$4^{+}$). $CD4^{+}$ and $CD8^{+}$ T-cell memory subsets were defined by differential expression of CD45RA, CCR7 and CD95 as in the resting panel.

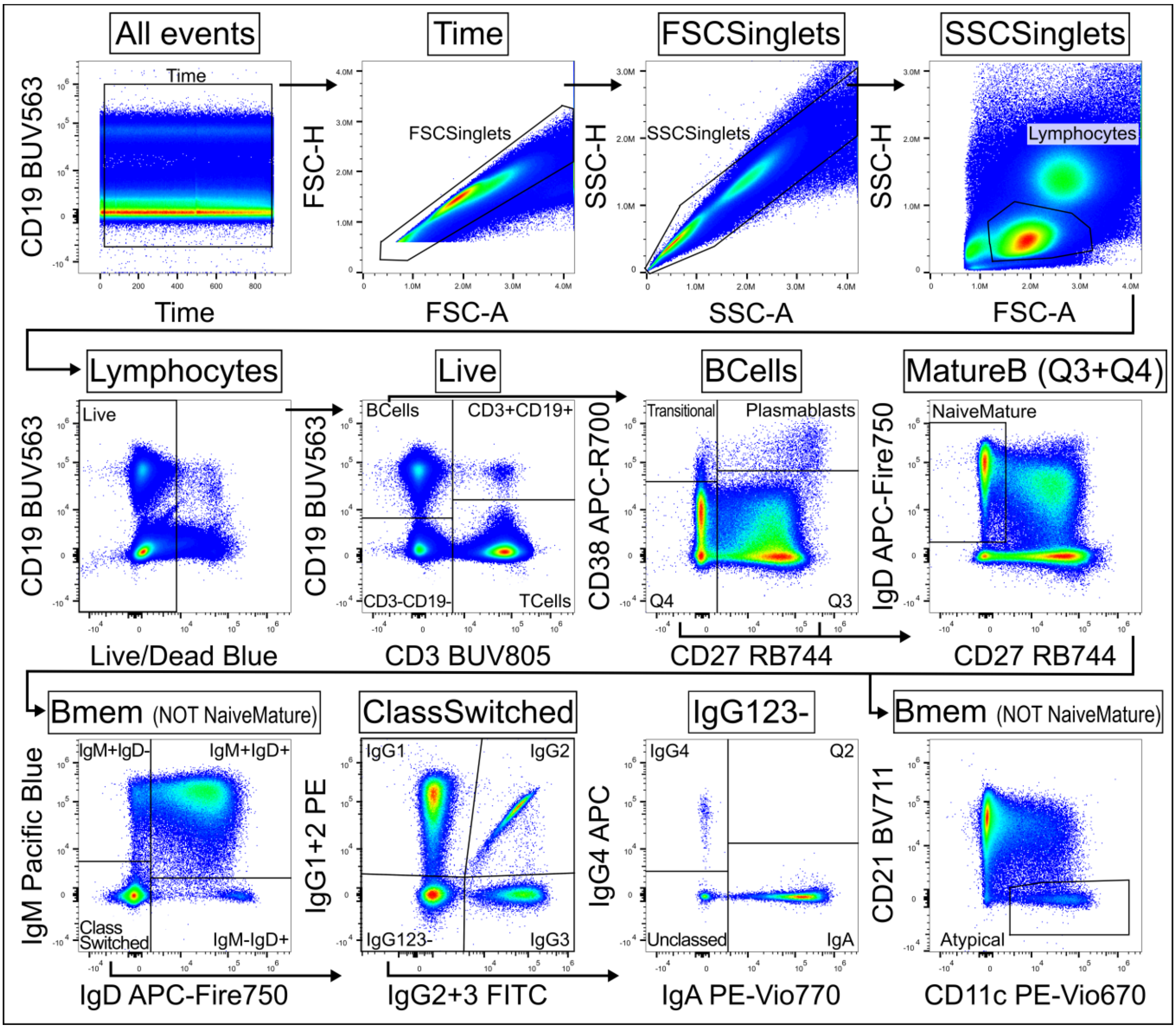


**Supplementary Figure 4.** Gating scheme for the 20-parameter B cell panel.

Events were gated sequentially to remove aggregates and select single, live lymphocytes (SSC$^{low}$FSC$^{mid}$Live/Dead Blue$^{-}$), before gating of CD19$^{+}$CD3$^{-}$ B cells. B cells were then gated to CD27$^{-}$CD38$^{high}$ transitional B cells and CD27$^{+}$CD38$^{high}$ plasmablasts, with the remainder of cells (Q3+Q4) representing mature B cells. Mature B cells were gated into CD27$^{-}$IgD$^{+}$ naive mature and memory B cells (Bmem; NOT naive mature). Bmem were then divided into IgM$^{+}$IgD$^{-}$, IgM$^{+}$IgD$^{+}$, IgM$^{-}$IgD$^{+}$, and class-switched, which were further gated to select IgG1$^{+}$, IgG2$^{+}$, IgG3$^{+}$, and from IgG1/2/3$^{-}$, IgG4$^{+}$ and IgA$^{+}$. Bmem cells not staining for any isotype were termed unclassified. Bmem were separately classed as CD21$^{-}$CD11c$^{+}$ atypical (atBC) and non-atypical.

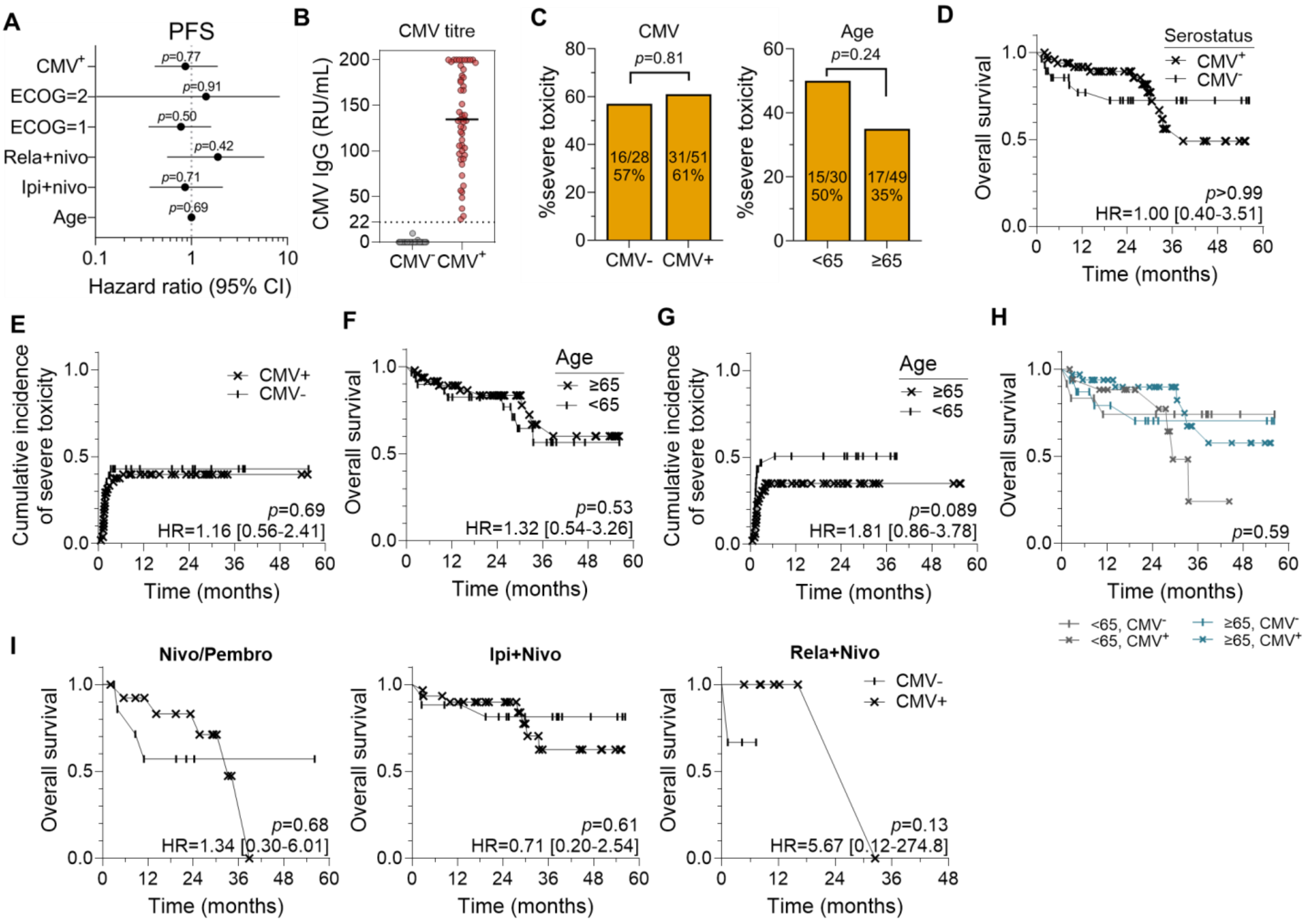


**Supplementary Figure 5: Clinical outcomes for advanced melanoma patients.**

(**A**) Cox proportional hazards model of CMV or age effect on progression-free survival (PFS) when controlling for ECOG performance status (reference = 0) and treatment (reference = nivo/pembro). Forest plot shows hazard ratio (HR) and 95% confidence interval (CI). (**B**) Anti-CMV capsid IgG titres. Cutoff for seropositivity was defined using kit controls at 22 relative units (RU)/mL. (**C**) Severe (grade 3+) toxicity rate when stratified by CMV serostatus or age. P values from Fisher's exact test. (**D**) Kaplan-Meier overall survival (OS) and (**E**) time to onset of severe toxicity when patients were stratified by CMV. (**F**) OS and (**G**) time to onset of high-grade toxicity in patients stratified by age. (**H**) Kaplan-Meier OS stratified by both age and CMV status. (**I**) Kaplan-Meier OS between CMV$^+$ and CMV$^-$ patients when assessed within each treatment cohort.

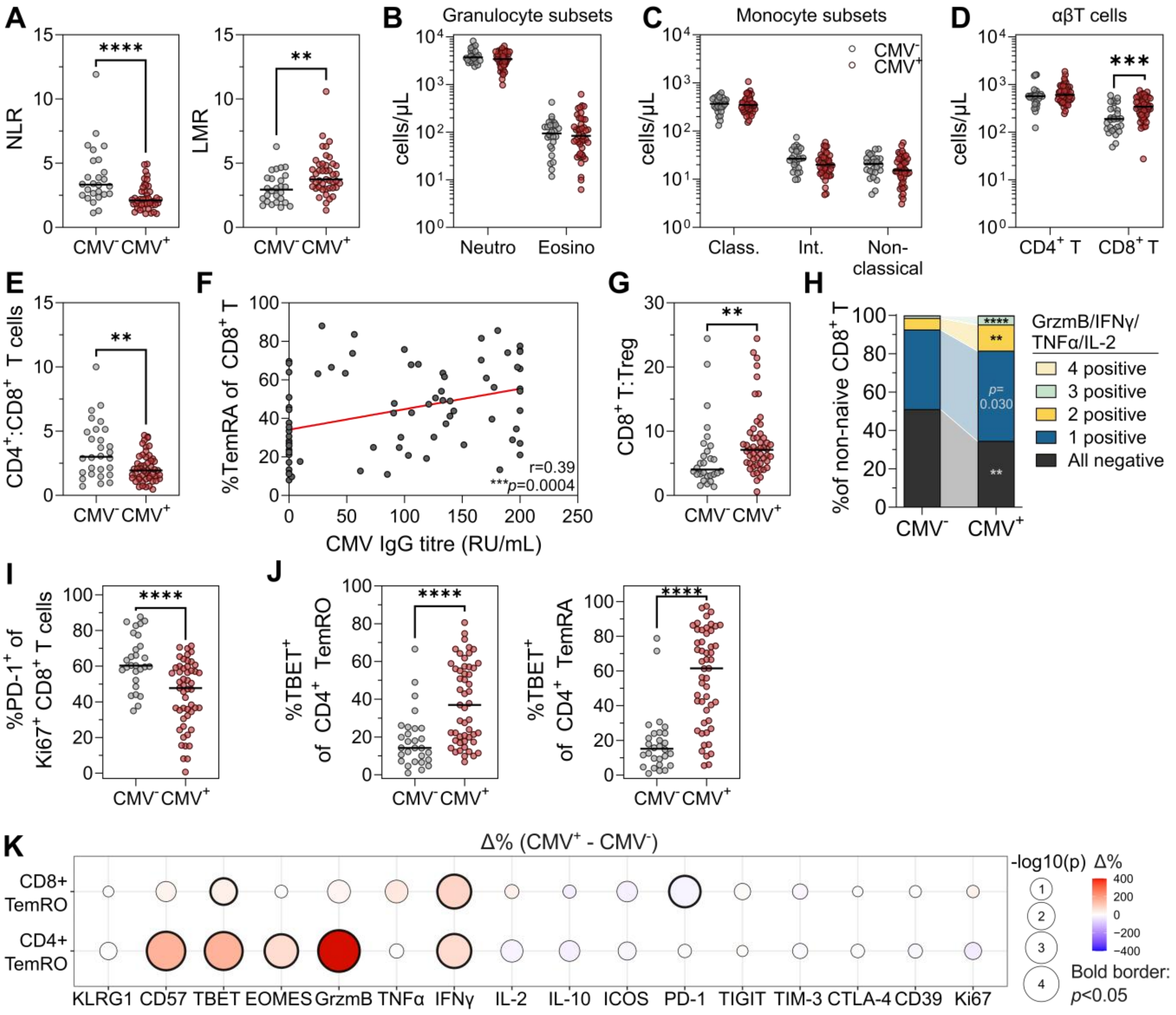


**Supplementary Figure 6: Pre-treatment immune phenotype in CMV⁻ and CMV⁺ patients.**

(**A**) Neutrophil to lymphocyte ratios (NLR) and lymphocyte to monocyte ratios (LMR) in CMV⁺ (n=51) and CMV⁻ patients (n=28). (**B**) Absolute counts of granulocyte (neutrophil, eosinophil) and (**C**) monocyte (classical, intermediate, and non-classical) subsets. (**D**) Absolute counts of CD8⁺ T cells. (**E**) CD4⁺ T to CD8⁺ T cell ratio. (**F**) Spearman correlation of proportional CD8⁺ TemRA cell abundance and anti-CMV capsid IgG titer. (**G**) CD8⁺ T to Treg cell ratio. (**H**) Proportion of CD8⁺ T cells expressing one or more of granzyme B, IFNγ, TNFα, and IL-2. (**I**) Proportion of proliferative Ki67⁺ CD8⁺ T cells expressing PD-1. (**J**) Proportion of CD4⁺ TemRO and TemRA cells expressing TBET. (**K**) Difference in proportion of CD8⁺ and CD4⁺ TemRO cells expressing markers between CMV⁻ and CMV⁺ patients. Balloon size represents -log10(p) value, with p-values < 0.05 marked by a bold border. Colour represents delta expression (CMV⁺ - CMV⁻); positive (red) values denote features upregulated in CMV⁺ patients. All plots show median values. *P* values from Mann-Whitney test; *$p<0.0167$, **$p<0.01$, ***$p<0.001$, ****$p<0.0001$.

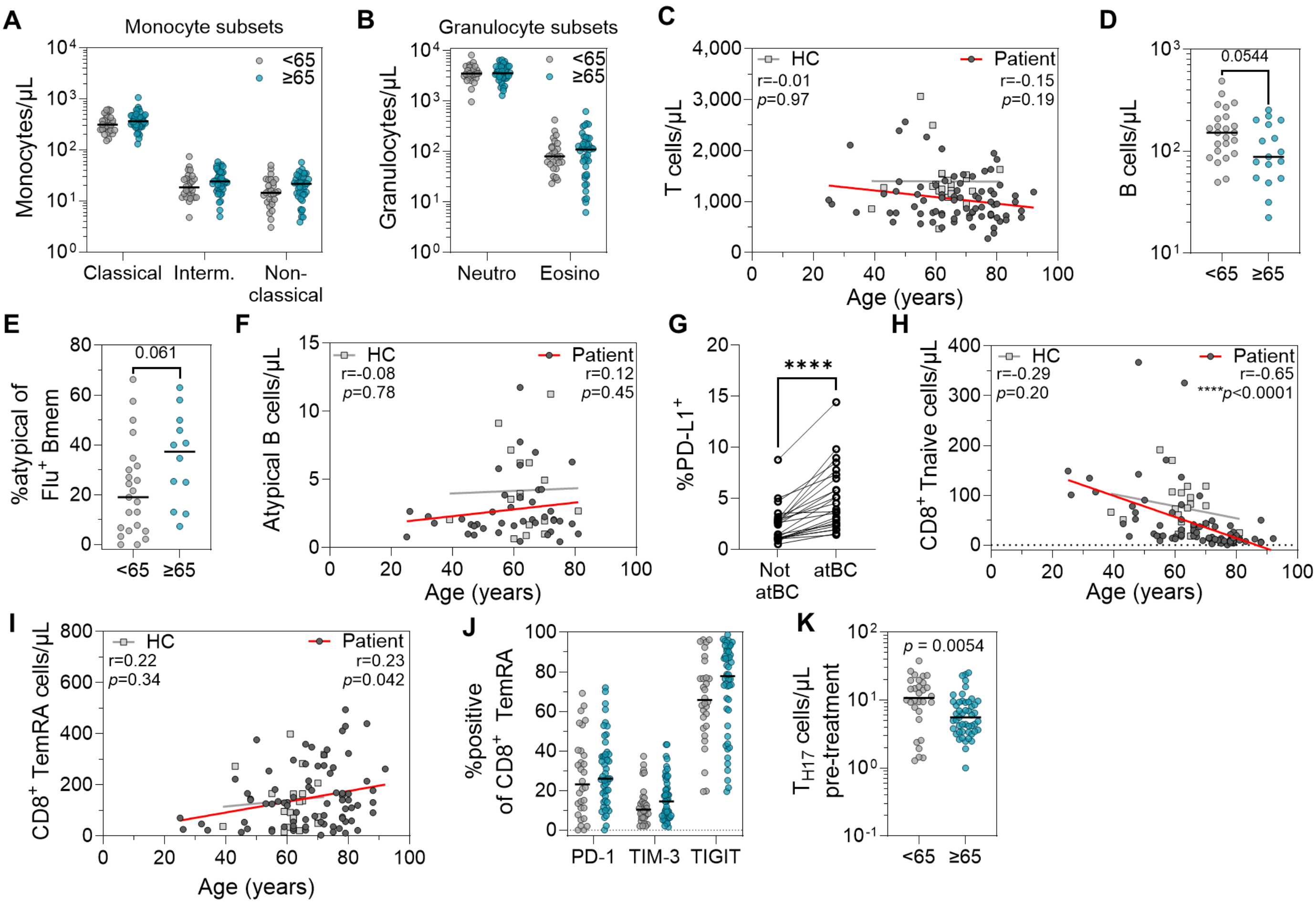


**Supplementary Figure 7**: **Pre-treatment phenotype in younger (<65 years old) and older (≥65) patients.** (**A**) Absolute counts of monocyte (classical, intermediate, and non-classical) and (**B**) granulocyte (neutrophil, eosinophil) subsets in younger (n=30) and older (n=49) patients. (**C**) Spearman correlation of absolute T cell abundance with age in patients (n=79) and healthy controls (HC; n=21). (**D**) Absolute B cell counts in ipi+nivo B cell flow cytometry cohort (n=41; 24 younger, 17 older). (**E**) Proportion of atypical B cells within influenza-specific memory B cell pool by age group. (**F**) Spearman correlation between absolute atypical B cell abundance and age in patients (n=41) and healthy controls (n=16). (**G**) PD-L1 expression frequency in atypical memory B cells (atBC) compared to conventional memory B cells (paired Wilcoxon test). (**H**) Spearman correlation between absolute $CD8^+$ T naive or (**I**) $CD8^+$ TemRA cell count and age for patients and healthy controls. (**J**) Frequency of PD-1, TIM-3, and TIGIT expression within $CD8^+$ TemRA cells in young and aged patients. (**K**) Absolute abundance of $T_{H17}$ cells. All plots show median values; *p* values from Mann Whitney tests unless otherwise noted. **p*<0.0167, ***p*<0.01, ****p*<0.001, *****p*<0.0001.

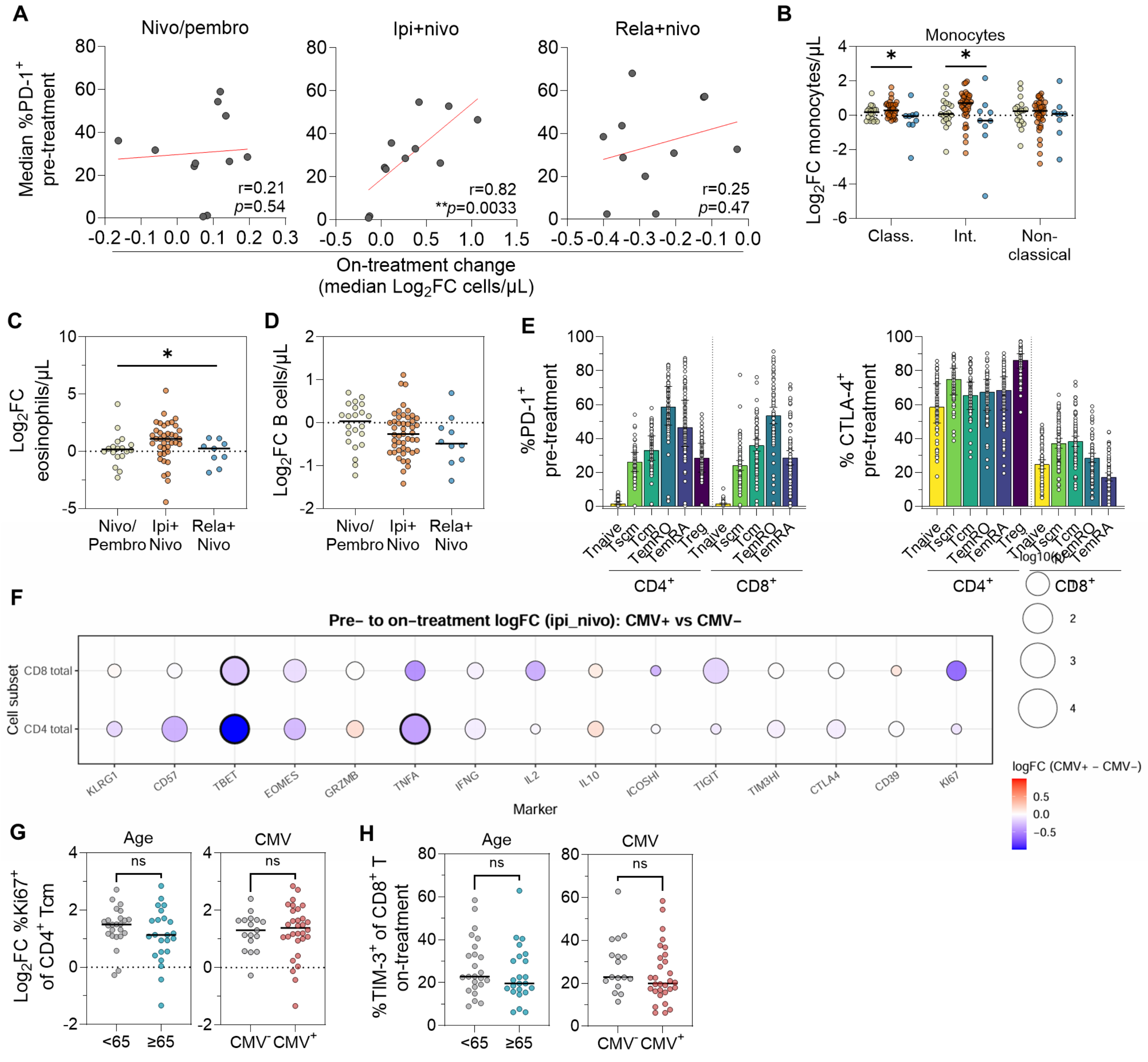


**Supplementary Figure 8: Immunological effects of nivo/pembro, ipi+nivo, and rela+nivo treatments.** (**A**) Spearman correlation between pre-treatment PD-1 expression frequency and on-treatment $\log_2$ fold change ($\text{Log}_2\text{FC}$) in absolute $CD4^+$ and $CD8^+$ T subset abundance by treatment. (**B**) Pre- to on-treatment $\text{Log}_2\text{FC}$ in classical (class.), intermediate (int.), and non-classical monocytes; (**C**) eosinophils, and (**D**) B cells. (**E**) Pre-treatment frequency of PD-1 and CTLA-4 expression per T cell subset. (**F**) Difference in pre- to on-treatment $\text{Log}_2\text{FC}$ in T cell marker expression frequency between $CMV^-$ and $CMV^+$ patients receiving ipi+nivo. Balloon size represents $p$ value; with $p$ values $< 0.05$ also marked by a bold border, while $\log_2\text{FC}$ difference is represented by colour. Positive (red) values represent features upregulated to a greater degree in $CMV^+$ patients. (**G**) Pre- to on-treatment $\log_2\text{FC}$ in Ki67 frequency within $CD4^+$ Tcm cells in ipi+nivo patients, stratified by age or CMV. (**H**) On-treatment TIM-3 expression frequency by $CD8^+$ T cells in ipi+nivo patients, stratified by age or CMV. All plots show median values. Statistical significance was assessed using Kruskal Wallis test (plots B-D), Mann Whitney tests (plots E, G, H).

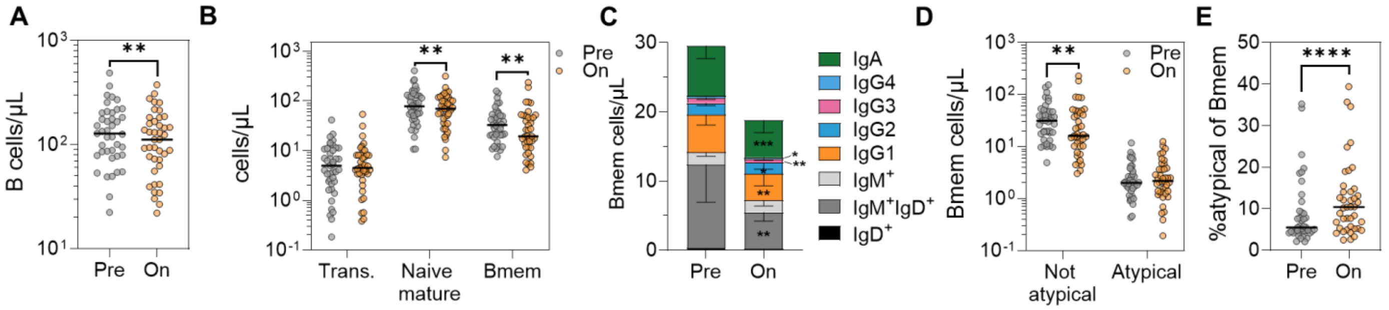


**Supplementary Figure 9: Effect of ipi+nivo treatment on B cells.**

(**A**) Absolute B cell count before and early during ipi+nivo treatment (n=41 paired). (**B**) Absolute counts of transitional (trans.), naive mature, and memory B cells (Bmem) pre- and on-treatment; and (**C**) abundance of Ig subclasses within Bmem. (**D**) Absolute and (**E**) proportional abundance of CD21$^{low}$ CD11c$^{+}$ atypical and non-atypical cells pre- and on-treatment. Plots show medians, *p* values from paired Wilcoxon tests.
